# SPEECH FEATURES IN A DEVELOPMENTAL PERSPECTIVE: A SCREENING TOOL FOR PSYCHOSIS, DEPRESSION AND ANXIETY

**DOI:** 10.64898/2026.10.28.26366635

**Authors:** Marina Ribeiro, Julia Amorim, Bárbara Malcorra, Renato Piccolo, Luísa Jensen, Diego Soares, Tamara Meletti, Natália B. Mota

## Abstract

Language production provides a window into cognitive development, social cognition, and mental health. To translate this potential into clinical practice, we developed a scalable speech-based platform integrating graph-theoretical, psycholinguistic, and emotional features from brief spontaneous narratives to support developmental monitoring, mental wellness screening, and clinical decision-making.

Normative developmental trajectories were derived from 2149 participants spanning 0–23 years of formal education. Narratives elicited using positively valenced pictures were transformed into word graphs to derive two composite indices: Narrative Complexity (NC), reflecting structural language organization, and Emotional Narrative Complexity (ENC), integrating narrative structure and emotional content. Both indices were modeled using asymptotic functions calibrated by schooling and converted into percentile-based normative curves. NC (R² = 0.29, τ = 7.77) and ENC (R² = 0.31, τ = 6.97) increased with schooling, approaching saturation around middle school. These trajectories provide educationally calibrated references for evaluating linguistic and socioemotional development.

Clinical utility was investigated in a transdiagnostic sample of 120 participants, comprising healthy controls (n = 38), mood disorders (n = 28), affective psychosis (n = 36), and non-affective psychosis (n = 18). Although ENC alone did not discriminate between symptom-risk or diagnostic groups, classifiers combining its constituent variables with education and sex achieved AUCs of 0.726 for symptom risk and 0.800 for diagnosis, increasing to 0.774 and 0.858, respectively, with additional linguistic features. Machine-learning models integrating graph-derived, psycholinguistic, and emotional features further estimated symptom dimensions assessed by PANSS, GAD-7, and PHQ-9, achieving AUCs of 0.74–0.92 across severity thresholds while maintaining sensitivity ≥ 0.80. Individual linguistic variables also differed significantly across symptom-severity strata, suggesting distinct associations with psychopathological dimensions.

Together, these findings establish an integrated speech-based framework spanning normative developmental assessment, mental wellness screening, and symptom-informed clinical support. By combining educationally calibrated trajectories with interpretable linguistic features and machine learning, this approach offers scalable digital markers to identify atypical developmental patterns, inform preventive strategies, and assist clinical assessment and decision-making.

## INTRODUCTION

Human language is one of the most complex forms of human behavior and provides a privileged window into cognitive, emotional, and social functioning. The way individuals use language reflects aspects of their thought organization, emotional states, and social behavior, providing a rich behavioral signal for studying human cognition and behavior (Mihalcea et al., 2024; Mota, Weissheimer, et al., 2023). Narrative production engages multiple cognitive processes involved in the expression of experiences, including lexical retrieval, working memory, executive control, temporal organization, and social and emotional processing (B. L. C. Malcorra et al., 2022; Mota, Weissheimer, et al., 2023).

Although oral language emerges naturally during development, the ability to produce coherent, hierarchically organized, and emotionally nuanced narratives is shaped by both developmental and environmental factors, including formal education and literacy experience (Mota, Ribeiro, et al., 2023; Mota, Weissheimer, et al., 2023). Schooling provides sustained exposure to increasingly complex linguistic structures, literacy practices, abstract concepts, and social interactions, which may contribute to the progressive refinement of communicative and narrative skills. Accordingly, narrative organization evolves across educational attainment and reflects the maturation of cognitive abilities as well as the acquisition and refinement of social-cognitive competencies involved in understanding, interpreting, and communicating experiences effectively (Gardner-Neblett, 2024; Grau-Husarikova et al., 2024; B. L. C. Malcorra et al., 2022; Mota et al., 2018, 2025; Mota, Weissheimer, et al., 2023; Suggate et al., 2018).

Emotional information constitutes a fundamental component of this developmental process. Beyond describing events, individuals must attribute emotional meaning to experiences, recognize affective states, and communicate them coherently and in a contextually appropriate manner. These abilities are closely related to social cognition and are essential for successful interpersonal functioning. Consequently, integrating emotional content into narratives may provide important insights into both cognitive development and emotional well-being (Grau-Husarikova et al., 2024; Mota, Ribeiro, et al., 2023). Understanding how emotional narrative abilities evolve across schooling is, therefore, critical for distinguishing typical developmental variation from potential indicators of psychological distress.

The developmental trajectory of language and social cognition unfolds within broader social and epidemiological contexts that also shape mental health. Social disadvantage, including poverty, educational deprivation, exposure to violence, and other forms of adversity, can constrain developmental opportunities and is associated with increased vulnerability to mental health problems across the life course (Lund et al., 2018; Marmot et al., 2012; Patel et al., 2008). Mental health conditions are among the leading causes of disability worldwide, and epidemiological studies estimate that around 30% of children and adolescents experience clinically relevant psychological difficulties (McGorry et al., 2024). These challenges highlight the need for scalable approaches that can monitor cognitive and emotional development across large populations while supporting early identification of individuals at risk.

Language-based assessment represents a promising strategy to address this need. Recent advances in computational psychiatry and natural language processing have demonstrated that spontaneous speech contains measurable markers of cognitive organization, emotional functioning, and psychopathology (Malgaroli et al., 2023; Palaniyappan, 2021). Automated analysis of speech has been successfully used to identify thought disorder, predict psychosis onset, characterize mood disorders, and monitor cognitive decline (Bedi et al., 2015; B. L. C. Malcorra et al., 2021; Morgan et al., 2021; Mota et al., 2012, 2014, 2017, 2025; Mota, Ribeiro, et al., 2023). Because speech can be collected quickly, remotely, and non-invasively, it represents an attractive candidate for large-scale screening and longitudinal monitoring.

Among computational approaches, graph-theoretical analysis of speech has emerged as one of the most robust and interpretable methods for quantifying thought organization (Mota et al., 2012, 2014, 2017, 2025; Mota, Ribeiro, et al., 2023). In this framework, words are represented as nodes and their sequential relationships as edges, generating mathematical representations of discourse structure (Mota et al., 2014; Mota, Weissheimer, et al., 2023). Graph attributes quantify dimensions such as lexical diversity, recurrence patterns, connectivity, and organizational complexity. Unlike semantic approaches that depend heavily on language-specific corpora and high-dimensional embeddings, graph-based methods capture structural properties of discourse that are largely independent of content, facilitating scalability and cross-cultural applicability (Mota, Weissheimer, et al., 2023). Over the past decade, graph measures of speech connectedness have consistently demonstrated sensitivity to psychosis, mood disorders, neurodevelopmental conditions, and cognitive impairment (Mota, Weissheimer, et al., 2023).

Despite these advances, an important challenge remains: language-based markers require developmental context for meaningful interpretation, as linguistic differences associated with educational experience may otherwise be confounded with psychopathology. Establishing normative developmental trajectories can therefore help distinguish expected variation from clinically relevant deviations. At the same time, the increasing use of complex machine-learning models in mental health raises concerns about clinical interpretability, highlighting the value of language-based markers that combine predictive performance with theoretically meaningful and clinically understandable features (Joyce et al., 2023).

The Mobile Brain framework was developed to address these complementary challenges. Building upon pioneering work on speech connectedness and graph-theoretical analysis of language (B. Malcorra et al., 2024; Mota et al., 2012, 2017, 2018, 2025; Mota, Ribeiro, et al., 2023), the framework combines graph-derived measures of discourse organization with psycholinguistic and emotional features extracted from brief spontaneous narratives. Rather than relying exclusively on black-box predictions, Mobile Brain generates interpretable indicators linked to meaningful dimensions of cognitive development, social cognition, emotional functioning, and mental health. The framework is designed to support a continuum of applications ranging from developmental monitoring and population-level screening to symptom-oriented clinical assessment.

The present study evaluates three complementary levels of language-based assessment. First, we characterize normative developmental trajectories of structural aspects of narrative and emotional expression across formal education using a large Brazilian sample spanning the full range of schooling, from individuals without formal education to highly educated adults. To achieve this objective, we introduce two composite language-based indices calibrated by years of formal education. The Narrative Complexity (NC) index captures structural aspects of language organization derived from graph attributes associated with lexical diversity, graph size, and short- and long-range recurrence patterns. The Emotional Narrative Complexity (ENC) index extends this framework by incorporating emotional expression and reflecting the integration of affective content into narrative organization. Our hypothesis is that both indexes develop according to formal education in an asymptotic trajectory, and must mature at around high school level, as observed in (Mota et al., 2018). Second, we investigate the utility of ENC as a scalable indicator of emotional well-being and mental health screening in a clinical and transdiagnostic population. We hypothesize that levels below the 25 percentile in the normative curve of ENC would be able to predict case-control participants in the clinical sample with sensibility higher than 80%. Third, we develop and validate machine-learning models that estimate symptom severity from spontaneous speech using clinically interpretable linguistic markers. The main hypothesis is that a specific combination of language markers would predict the risk of psychotic, depressive and anxious symptoms measured by psychometric scales. Together, these analyses establish a unified framework spanning developmental assessment, mental wellness screening, and symptom-informed clinical decision support, providing a practical pathway from large-scale monitoring to individualized mental health evaluation via a brief, scalable speech-elicitation protocol.

## METHODS

This study consisted of two cohorts: one spontaneously collected by Mobile Brain users (study approved by the ethics committee - CAAE n° 91594425.8.0000.5282 with de-identified data), hereby referred as the large-scale cohort, with a broad educational attainment and age range, and the other is a clinical sample from mental health specialized centers (study approved by the ethics committee - CAAE n° 86286024.5.1001.5263 with participant-consented personal data), hereby referred as the symptoms cohort. The collection, processing and analysis of these datasets were done as described in the following sections.

### 1. Large-scale and symptom datasets description

The large-scale dataset comprised 2149 participants, spanning a broad developmental spectrum from early childhood to adulthood. The participants used Mobile Brain Platform as consumers of educational or mental health screening products by Mobile Brain from different projects around the country (Brazil). Participants had a mean educational attainment of 6.40 years of formal schooling (SD = 4.78; range = 0–23 years), including individuals with no formal education and those with advanced educational backgrounds. Regarding biological sex, 669 participants (31.1%) were classified as female, 719 (33.5%) as male, and 761 (35.4%) had no sex information available. Among participants with identified sex (n = 1388), 48.2% were female, and 51.8% were male. Mean years of education were comparable between female (mean = 6.95, SD = 5.44) and male participants (mean = 7.00, SD = 5.29).

The second dataset (symptoms dataset) consists of individual-level observations, incorporating spontaneous speech samples alongside demographic and clinical parameters. The sample includes 130 data points derived from 120 unique individuals (58 females and 62 males). All participants were clinically evaluated by a mental health specialist, and diagnostic criteria from DSM V were used to categorized groups as follows: healthy controls (n = 38), individuals with mood or anxiety disorders (n = 28), affective psychosis like bipolar disorders (n = 36), and non-affective psychosis like schizophrenia (n = 18). Their ages ranged from 18 to 79 years (mean 41.5 years), and their educational attainment ranged from 2 to 22 years of formal schooling (mean 12.7 years).

### 2. Speech elicitation protocol and text preprocessing

Speech samples were collected using the Happy Thoughts protocol (Mota, Ribeiro, et al., 2023). The protocol consists of the sequential presentation of three positively valenced images. For each stimulus, participants were instructed to observe the image for 15 seconds. After the image disappeared from the screen, they were asked to tell a story about what they had seen. This procedure was repeated for all three images within the same assessment session. The stimuli depicted three thematic categories (babies, dogs, and desserts) selected from the International Affective Picture System (IAPS) (Lang et al., 2020) due to their positive affective valence. To minimize order effects, the image presentation sequence was randomized across sessions. Beyond the Happy Thoughts protocol, symptom participants afterward answered psychometric scales GAD-7 and PHQ-9, followed by an application of the PANSS psychometric scale by a trained mental health specialist (exclusively for the second sample of participants).

Audio recordings were de-identified prior to analysis and automatically transcribed using the *Whisper* large language model developed by *OpenAI* (Radford et al., 2022). The transcriptions of the three narratives elicited during a session were concatenated into a single text, which served as input for subsequent computational analyses. This procedure allowed the extraction of multiple linguistic indicators from a standardized speech sample while preserving the ecological validity of spontaneous narrative production. The Happy Thoughts protocol, as well as the mental health questionnaires, de-identification and preprocessing of recordings were implemented and run end-to-end automatically through a web-based application developed by Mobile Brain S.A.

### 3. Speech markers

Narratives were converted into directed word graphs in which each unique word represented a node and the temporal succession between words was represented by edges. From these graphs, structural attributes related to lexical diversity, short-range recurrences, long-range recurrences, and graph size were computed.

Graph-theoretical analyses were performed using a proprietary algorithm designed to generate word recurrence graphs. In these directed graphs, nodes represent unique words, and edges represent the temporal sequence connecting consecutive words throughout the narrative. Because graph measures are sensitive to differences in text length and participants naturally vary in speech verbosity, graph attributes were calculated using a moving-window approach. Specifically, graphs were constructed from windows containing 30 words, with steps of 15 words, guaranteeing 50% of overlap between consecutive graphs. Final graph metrics were obtained by averaging the measures across all windows within each narrative sample. The complete list of graph attributes is provided in Table 1.

**Table 1.** Graph features extracted from narrative speech. This table presents the topological attributes calculated using 30-word moving windows, including metrics for connectivity, network size, and short-range recurrences. NA - Data not available

| Graph structure features |  |  |  |
| --- | --- | --- | --- |
| Variable | Description | Values (symptom) | Values (large-scale) |
| N | Mean of the number of nodes calculated with graphs of 30 words windows | mean 24.65, sd 1.28 | mean 23.56, sd 2.27 |
| E | Mean of the number of edges calculated with graphs of 30 words windows | mean 28.62, sd 0.22 | NA |
| ASP | Mean of the average shortest path calculated with graphs of 30 words windows | mean 4.59, sd 0.52 | mean 4.22, sd 0.75 |
| LSC | Mean of the largest strongly connected component calculated with graphs of 30 words windows | mean 16.09, sd 1.91 | mean 15.01, sd 3.92 |
| LCC | Mean of the largest connected component calculated with graphs of 30 words windows | mean 23.94, sd 1.32 | NA |
| RE | Mean of the repeated edges calculated with graphs of 30 words windows | mean 0.92, sd 0.68 | mean 1.39, sd 1.54 |

Additional linguistic features were extracted using *Stanza’s* natural language processing toolkit (Qi et al., 2020) implemented in Python 3.11. Universal Dependencies parsing was employed to derive morphosyntactic and syntactic indicators from the transcribed narratives. Emotional valence was estimated using a BERT-based language model pretrained on a proprietary Mobile Brain dataset of spontaneous speech and fine-tuned to estimate continuous positive- and negative-emotion scores (Devlin et al., 2018). The verbosity of speech was assessed using the *long_words* metric, which represents the density of words exceeding 7 characters. Finally, narrative adherence to the elicited stimuli (*on-topic* speech) was assessed by computing the cosine similarity between sentence embeddings from participants’ narratives and artificial-intelligence-generated descriptions of the specific images presented during the protocol. Embeddings were obtained using the *Sentence-BERT* framework (Reimers & Gurevych, 2019), employing a multilingual architecture (Reimers & Gurevych, 2020). Higher similarity scores indicated greater semantic alignment between the produced narratives and the eliciting visual stimuli. A comprehensive synthesis of demographic, clinical, structural, and semantic variables is provided in Supplementary Table 1.

### 4. Statistical modeling of educational attainment and psychiatric symptoms

#### a. Large-scale sample and construction of developmental curves

Based on previous studies demonstrating their association with cognitive maturation and formal education (Mota et al., 2018; Pinheiro et al., 2020), the graph-derived measures were combined in a linear regression, defined as Narrative Complexity (NC) index, calibrated by years of formal education. Higher NC values indicate increasingly organized and developmentally mature narrative structures.

To account for the affective dimension of discourse, emotional valence information derived from the narratives was incorporated into the structural model, yielding the Emotional Narrative Complexity (ENC) index. It calculated the proportion of positive and negative emotional words used in the narrative. As the narrative were based on positive affective pictures, the proportion of positive emotional words were considered as a congruent emotional expression, and the proportion of negative emotional words were considered as an incongruent emotional expression, revealing negative bias. Both proportions were combined with narrative graph attributes in a linear regression calibrated by years of formal education.

To establish normative developmental trajectories, NC and ENC scores were modeled as a function of years of formal education using an asymptotic exponential growth function:

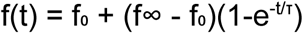

where (f₀) represents the initial value of the index, (f∞) represents the asymptotic maximum value reached at developmental saturation, (t) corresponds to years of formal education, and (τ) is the characteristic time constant of the trajectory. The parameter (τ) quantifies the educational pace of development, indicating how rapidly narrative abilities approach their asymptotic level as schooling increases. This function is the solution of the first-order differential equation:

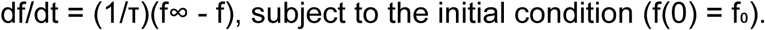

This modeling approach was selected based on the ontogenetic assumption that cognitive development follows nonlinear growth dynamics characterized by periods of rapid change followed by progressive stabilization (Mota et al., 2018; Pinheiro et al., 2020). Model parameters were estimated from the normative sample, yielding fitted developmental curves that represent the expected maturation of narrative complexity and emotional narrative organization across educational levels. The fitted functions were subsequently used to generate percentile-based normative curves, allowing individual scores to be positioned relative to the expected developmental trajectory for a given level of schooling.

Finally, percentile bands were derived from the fitted models to create normative developmental curves for both NC and ENC. These percentile curves function analogously to pediatric growth charts, allowing individual performance to be compared against expected values for a given educational stage. The resulting reference framework enables the identification of individuals presenting narrative development below, within, or above the expected range for their level of schooling. It supports the use of these indices as large-scale screening tools in typically developing populations.

Having established the normative developmental trajectories of narrative and emotional complexity across formal education, the subsequent objective was to evaluate the clinical utility of these linguistic markers. To determine whether speech-derived metrics can identify deviations associated with psychological distress, we applied the *Mobile Brain* framework to a transdiagnostic psychiatric cohort. The following analyses investigate the efficacy of these structural and emotional speech features in screening for general mental wellness, distinguishing among diagnostic groups, and mapping specific symptom-severity dimensions, including anxiety, depression, and psychosis, onto continuous language parameters.

#### b. Assessing the triage performance of the Emotional Narrative Complexity index

Leveraging the clinical dataset, we evaluated the discriminative capacity of the Emotional Narrative Complexity index to effectively stratify participants into distinct clinical categories. This was done using a case-control framework to distinguish healthy controls from individuals with any psychiatric diagnosis present within the cohort. To ensure statistical equivalence in comparative analyses, the clinical subgroup underwent a random downsampling procedure, repeated 10 times, to match the control cohort’s dimensionality. The final predictive performance of the linguistic indicator was subsequently determined by calculating the mean classification accuracy across these 10 balanced iterations. We also tested a risk of symptoms outcome: the risk group is defined by a participant with a score above any of the clinical scale cutoffs: 10 points for PHQ-9, 10 points for GAD-7, 14 points for PANSS positive, 14 points for PANSS negative or 60 points for PANSS total.

#### c. Symptom-level modeling of psychiatric scales

At the symptom level, the discriminative power of individual language markers was first assessed using Spearman’s rank correlation analyses between speech-derived metrics and standardized clinical scores, with Benjamini-Hochberg corrections for multiple comparisons. Furthermore, to simulate a population-level screening framework, we binarized psychometric outcomes using three threshold benchmarks: a permissive criterion designed to optimize screening sensitivity, a conservative parameter intended to equilibrate specificity and sensitivity (Pellas & Damberg, 2021; Plummer et al., 2016), and the conventional clinical cutoff. Because the PANSS was primarily designed to monitor symptom severity in clinical populations rather than as a primary screening instrument, we established triage benchmarks based on literature-derived criteria for clinical remission and psychological well-being (Lasser et al., 2007; Leucht et al., 2005). A summary of the chosen cutoffs is shown in Table 3.

**Table 3.**
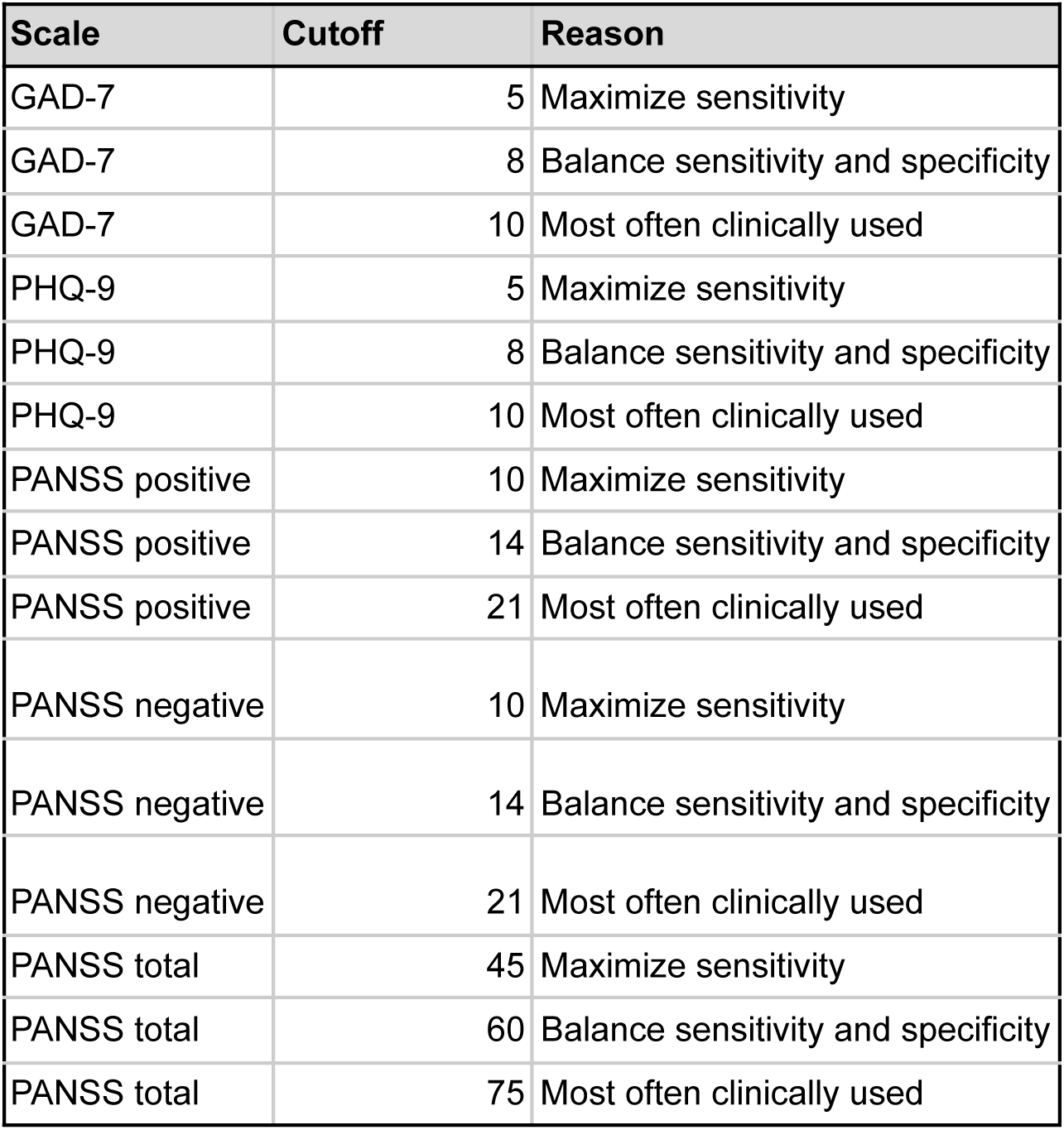
Diagnostic threshold benchmarks for clinical scale binarization. The table outlines the specific cutoffs applied to the GAD-7, PHQ-9, and PANSS instruments, stratified by their screening objective: maximizing diagnostic sensitivity, balancing sensitivity and specificity, or reflecting conventional clinical practice.

To evaluate the predictive utility of language-based markers, we developed a classification framework to identify symptom-severity dimensions with high sensitivity. We implemented a correlation-based network analysis using the *qgraph* framework in R (Epskamp et al., 2012) to reduce dimensionality. Initially, Pearson correlation coefficients were computed across a set of 37 structural, lexical, and syntactic features. These variables were then visualized using a force-directed layout based on the Fruchterman-Reingold algorithm, enabling topological inspection of feature clusters and identification of redundant metrics. Based on these observed aggregations, we derived composite indices by linearly combining correlated variables. The formulas for these integrated indicators are detailed in Table 5.

Using these integrated indices, we implemented a supervised machine-learning architecture designed for clinical screening applications. Initially, continuous variables were standardized as z-scores to ensure uniform scaling across diverse linguistic indicators. Predictive modeling was conducted through Bayesian logistic regression via the *rstanarm* framework, targeting the binary classification of symptom severity (Normal vs Risk) across all clinical outcomes and diagnostic thresholds. To mitigate overfitting and facilitate feature selection, we employed a Bayesian LASSO prior for coefficient estimation. The modeling strategy included interaction terms between composite speech indices and biological sex, while adjusting for potential confounders, including years of formal education. To enhance the framework’s effectiveness as a screening instrument, decision boundaries were determined using the pROC package and Receiver Operating Characteristic (ROC) analysis. We adopted a constrained optimization procedure that prioritized diagnostic sensitivity over standard default parameters; specifically, thresholds were calibrated to maximize specificity while maintaining a mandatory sensitivity floor of 80% (Bujang, 2026). The model’s performance was quantified using Area Under the Curve (AUC), optimized sensitivity and specificity metrics, and confusion matrix distributions for each psychiatric dimension.

**Figure 1.**
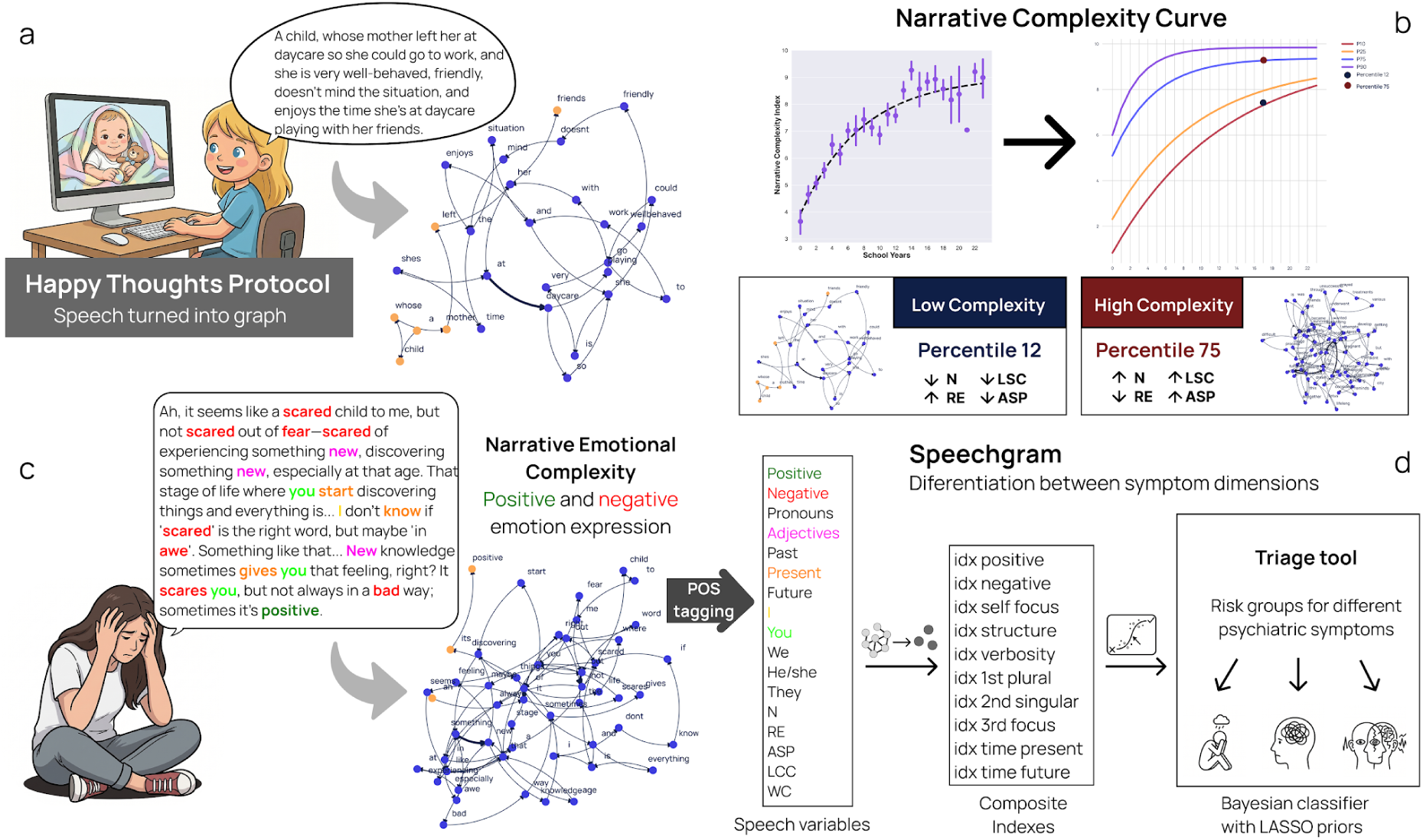
Overview of the *Mobile Brain* methodological pipeline for extracting speech features. **(a)** Application of the *Happy Thoughts* protocol, where spontaneous narratives are elicited from positive images and converted into directed word graphs. **(b)** Extraction of the Narrative Complexity (NC) index, illustrating the developmental curve alongside topological examples of low-(12th percentile) and high-complexity (75th percentile) graphs. **(c)** Extraction of the Emotional Narrative Complexity (ENC) index, demonstrating Part-of-Speech (POS) tagging and the incorporation of positive and negative emotional valence into the narrative structure. **(d)** Generation of the *Speechgram* by grouping speech variables into composite indices (e.g., temporal focus, structure, verbosity), which feed a Bayesian classifier functioning as a triage tool for different psychiatric dimensions.

## RESULTS

The resulting Narrative Complexity (NC) index was positively associated with years of formal education (Pearson-r = 0.516, R² = 0.266, p << 0.001), indicating progressively more organized and developmentally mature narrative structures with increasing educational attainment. Incorporating emotional valence into the structural model yielded the Emotional Narrative Complexity (ENC) index, which showed a similar positive association with years of formal education (Pearson-r = 0.530, R² = 0.281, p << 0.001). Given the positive valence of the eliciting stimuli, positive emotional language represented congruent emotional expression, whereas negative emotional language represented incongruent expression and negative affective bias. Together, these findings indicate that both structural and affective dimensions of spontaneous narratives vary systematically with educational attainment, supporting the subsequent modeling of NC and ENC normative developmental trajectories.

### 1. Asymptotic Growth of Narrative and Emotional Narrative Complexity as a Function of Educational Attainment

Narrative Complexity (NC) and Emotional Narrative Complexity (ENC) increased with educational attainment, consistent with the expected asymptotic developmental pattern described by the fitted exponential growth model. In both cases, rapid gains were observed during the early years of schooling, followed by a progressive deceleration toward a plateau in later educational stages.

For Narrative Complexity, the asymptotic model provided a significant fit to the data (R² = 0.29, MSE = 4.33). The estimated initial value was f₀ = 3.93, corresponding to the beginning of formal literacy acquisition (approximately 4 years of schooling), while the asymptotic value reached f∞ = 9.04. The characteristic time parameter was τ = 7.77 years of schooling, indicating that most developmental gains occurred by approximately the eighth year of education, corresponding to the final years of Brazilian middle school. After this point, increases in Narrative Complexity became progressively smaller, suggesting a saturation process in the maturation of discourse organization.

A similar developmental trajectory was observed for Emotional Narrative Complexity. The fitted model explained a comparable proportion of the variance (R² = 0.31, MSE = 4.45), with an estimated initial value of f₀ = 3.69 and an asymptotic value of f∞ = 8.94. The characteristic time parameter (τ = 6.97 years of schooling) indicated a slightly faster developmental trajectory than that observed for Narrative Complexity, with emotional narrative organization approaching its asymptotic range around the seventh year of schooling (also during middle school). As observed for NC, the growth rate decreased substantially after the elementary school years and approached a stable plateau during later educational stages.

**Figure 2.**
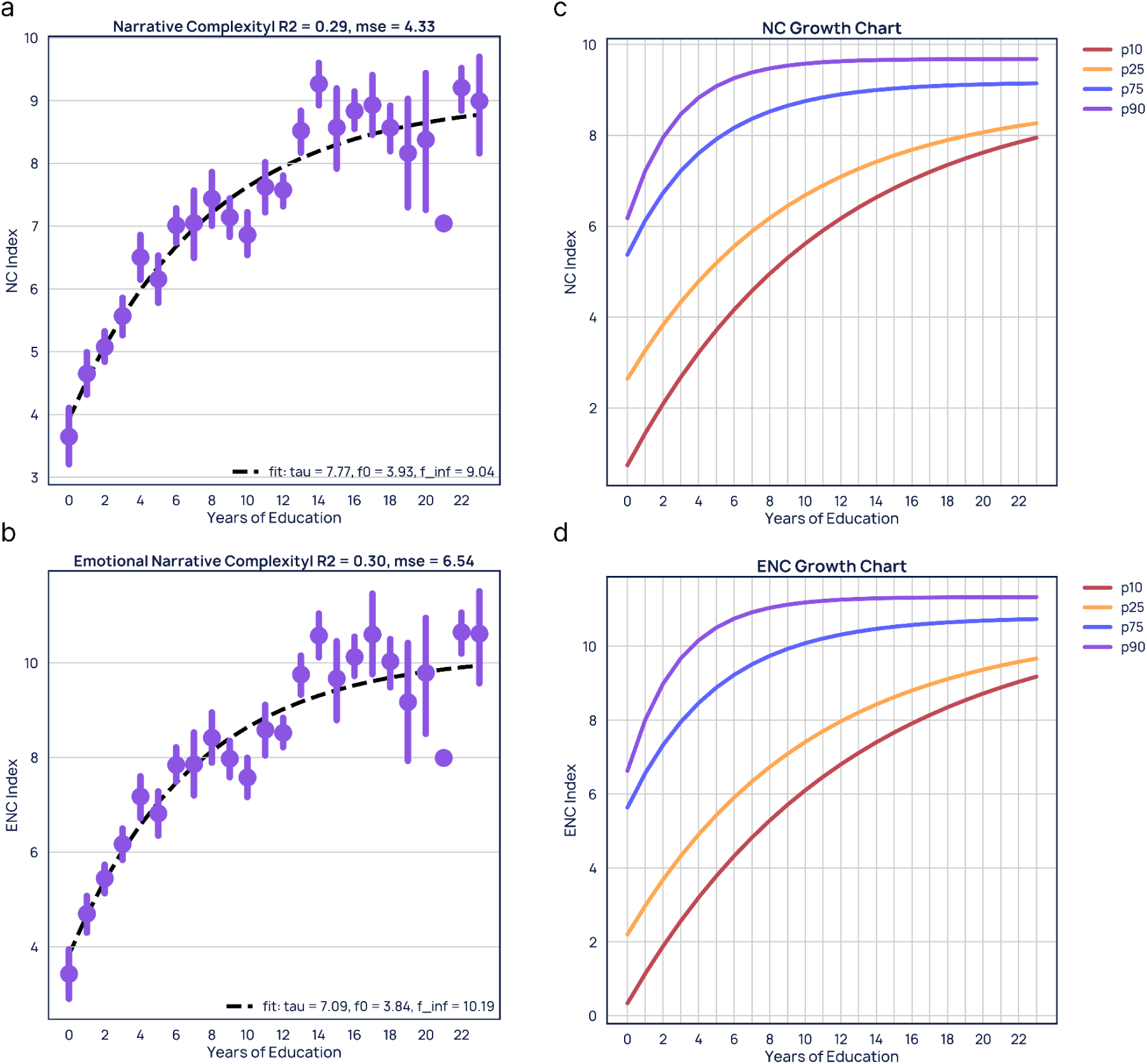
Developmental trajectories and normative curves as a function of formal educational attainment (years of schooling). **(a)** Asymptotic exponential growth model fit for the Narrative Complexity (NC) index. **(b)** Corresponding asymptotic exponential growth model fit for the Emotional Narrative Complexity (ENC) index. **(c)** Percentile-based normative growth curves (p10, p25, p75, p90) for the NC index, serving as expected developmental references across educational stages. **(d)** Percentile-based normative growth curves for the ENC index.

Based on these fitted trajectories, percentile-based normative curves were generated for both measures across the entire educational range represented in the sample. These curves provide educationally calibrated reference values that allow individual performance to be positioned relative to a large normative population spanning from individuals without formal schooling to highly educated adults. Consequently, NC and ENC can be interpreted as developmental indicators, analogous to growth charts used in physical health, enabling the identification of individuals performing below, within, or above the expected range for their level of educational attainment.

Overall, the results indicate that both structural and emotional dimensions of narrative production exhibit systematic developmental growth across schooling, with the most pronounced gains occurring during the years immediately following literacy acquisition and approaching asymptotic stabilization by the end of elementary education. These findings support the use of NC and ENC as normative markers of cognitive and emotional development in typical populations.

### 2. Predictive power of ENC as screening tool for mental wellbeing

Having established this normative developmental baseline, we sought to determine if deviations in the Emotional Narrative Complexity (ENC) index could serve as an initial marker for psychological distress. Neither the ENC index, nor its percentile values based on the development curve proved to be a viable indicator for general clinical assessment, either for a case-control framework or as a risk of symptoms (Table 4). The low performance observed in this scenario indicates that despite being an easy-to-implement, simple radar for mental wellness, this framework is limited to assess the normative development of speech, and other complimentary tools are needed to understand the linguistic features of mental suffering.

**Table 4.** Discriminative performance of the Emotional Narrative Complexity (ENC) index as a clinical indicator. The table presents the index’s standalone capacity to classify participants regarding mental wellness (individuals without clinical risk) and in a case-control scenario (healthy controls vs. psychiatric disorders). Performance is reported using the Area Under the ROC Curve (AUC), Sensitivity, and Specificity.

| Outcome | AUC | Sensitivity | Specificity |
| --- | --- | --- | --- |
| Risk of symptoms | 0.438 | 0.926 | 0.06 |
| Case-control (diagnosis) | 0.466 | 0.826 | 0.137 |

### 3. Individual speech variables are distinct between symptom severity levels

To achieve symptom-level specificity, we first evaluated the linear and monotonic relationships between continuous speech-derived metrics and standardized clinical scores using Spearman correlations (Figure 3). Some cross-diagnostic patterns emerged across multiple dimensions.

**Figure 3.**
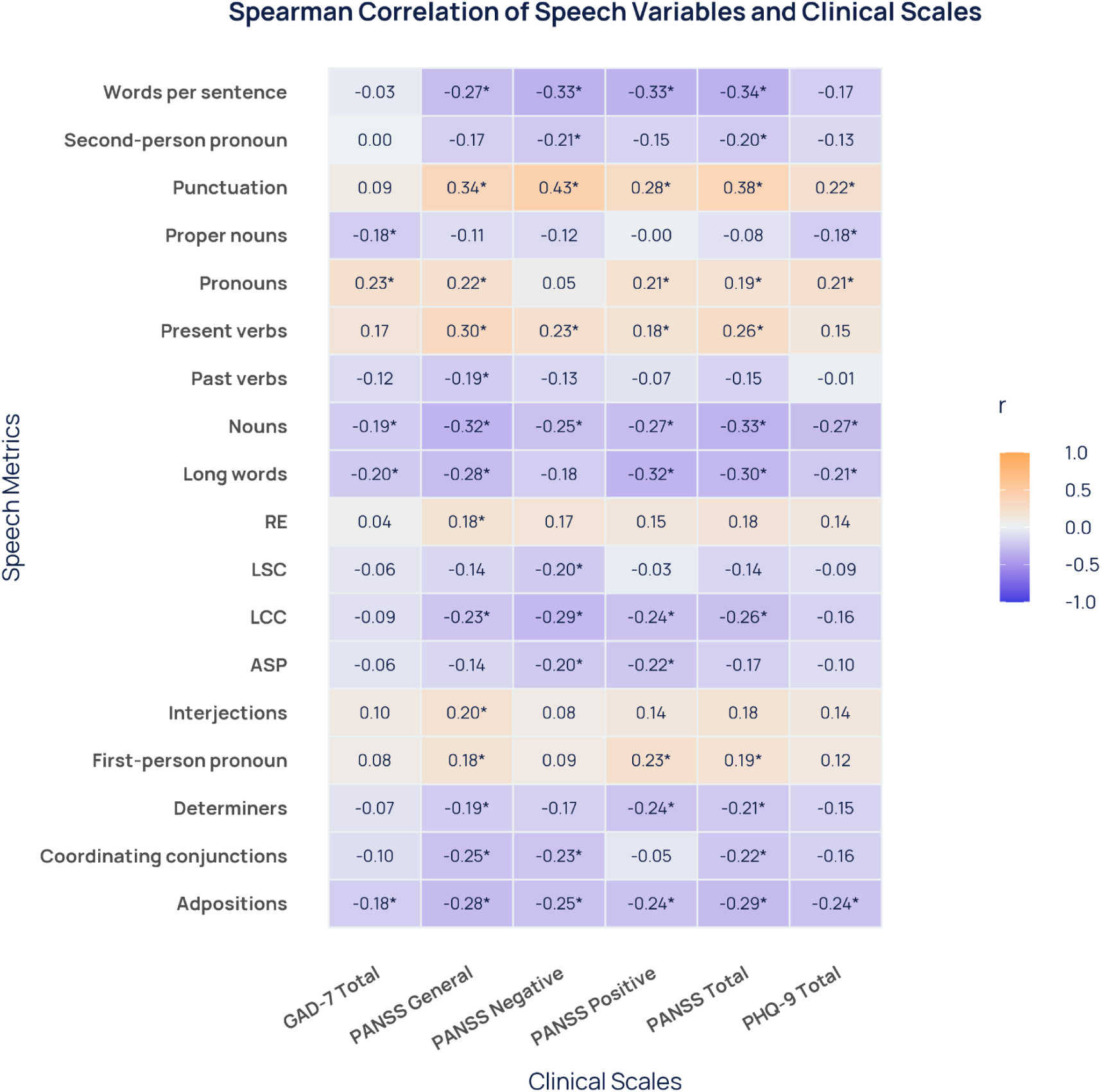
Spearman correlation matrix evaluating the monotonic relationships between a broad set of continuous speech metrics (e.g., words per sentence, punctuations, word classes) and standardized clinical scale scores. Significant correlation (Benjamini-Hochberg adjusted p-values below 0.05) tiles are marked with a * symbol on the side of the value. Rows with no significant correlations were omitted

Greater anxiety symptom severity (GAD-7) was associated with lower frequencies of nouns, proper nouns, and adpositions, shorter words, and a higher frequency of pronouns. A similar pattern was observed for depressive symptom severity (PHQ-9), with the additional finding that greater severity was associated with more frequent punctuation (Figure 3).

Psychotic symptoms showed a broader pattern of linguistic associations. Greater overall psychotic symptom severity (PANSS total score), as well as greater severity across all three PANSS subscales, was associated with fewer words per sentence, lower noun and adposition frequencies, smaller largest connected components (LCCs) in the word recurrence graphs, more frequent punctuation, and greater use of the present tense (Figure 3).

Despite these shared patterns, symptom-specific associations were also observed. Greater severity of negative symptoms was uniquely associated with a lower frequency of second-person pronouns and smaller LSC and ASP values (Figure 3). In contrast, greater severity of positive and general symptoms was associated with higher pronoun frequencies, particularly first-person pronouns, as well as shorter words and lower determiner frequencies (Figure 3).

These findings suggest that individual speech variables do not merely drift uniformly under generalized stress, but rather break down into granular, distinct structural patterns that are highly specific to individual psychiatric symptom dimensions, and that this can be used to differentiate mental suffering into symptom categories.

### 4. A Bayesian classifier with compounded linguistic indexes as an efficient triage tool for distinguishing between symptoms

To map the numerous speech features to clinical outcomes while reducing the model’s complexity, we first performed a correlation-based network analysis (Figure 4b). The force-directed layout revealed clusters grouping redundant linguistic metrics among the initial 37 variables. Based on these mathematical aggregations, ten distinct composite indicators were derived via linear combination to capture overarching linguistic dimensions (Table 5 and Figure 4b). For instance, structural markers were consolidated into idx_structure, incorporating Largest Connected Component (LCC), Average Shortest Path (ASP), Number of nodes (N), and Repeated Edges (RE) (Table 5 and Figure 4b). At the same time, production and length variations were captured by the idx_verbosity metrics: Word Count (WC), Edges (E), Largest Strongly Connected Component (LSC), and Words per Sentence (WPS) (Table 5 and Figure 4b). Self-referential patterns and temporal orientations were similarly isolated into targeted indices, such as idx_self_focus, idx_time_present, and idx_time_future, thereby providing a simplified yet detailed architecture for the model (Table 5 and Figure 4b). Emotional patterns emerged such as idx_positive combining positive emotional and adjectives, and idx_negative with negative emotional words isolately (Table 5 and Figure 4b). An index with a cluster of grammatical classes but an unrelated lexical function was observed but omitted from the model construction to maintain clinical explainability.

**Figure 4.**
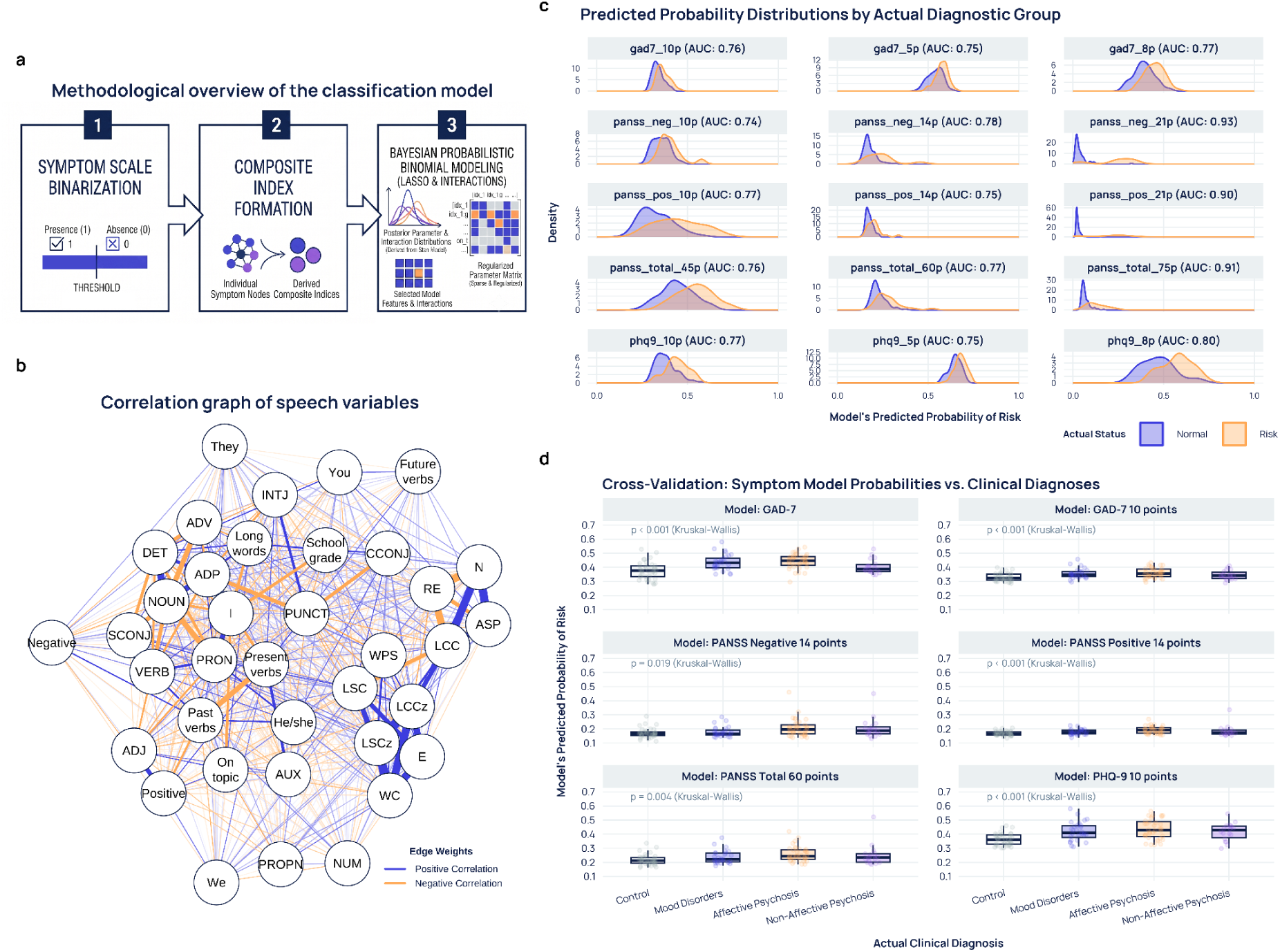
Structure and performance evaluation of the predictive architecture for screening symptom severity. **(a)** Methodological scheme of the classifier, comprising symptom scale binarization, composite index formation, and Bayesian binomial modeling with LASSO priors. **(b)** Force-directed network graph based on Pearson correlations among 37 speech variables, used to identify redundant clusters and guide dimensionality reduction. **(c)** Density plots illustrating the model’s predicted probability distributions of risk versus the actual status (Normal vs. Risk), highlighting discrimination across distinct clinical thresholds for the GAD-7, PANSS, and PHQ-9 scales. **(d)** Boxplots displaying the predicted probability of risk against actual clinical diagnoses in cross-validation (Controls, Mood Disorders, Affective Psychosis, and Non-Affective Psychosis).

**Table 5.**
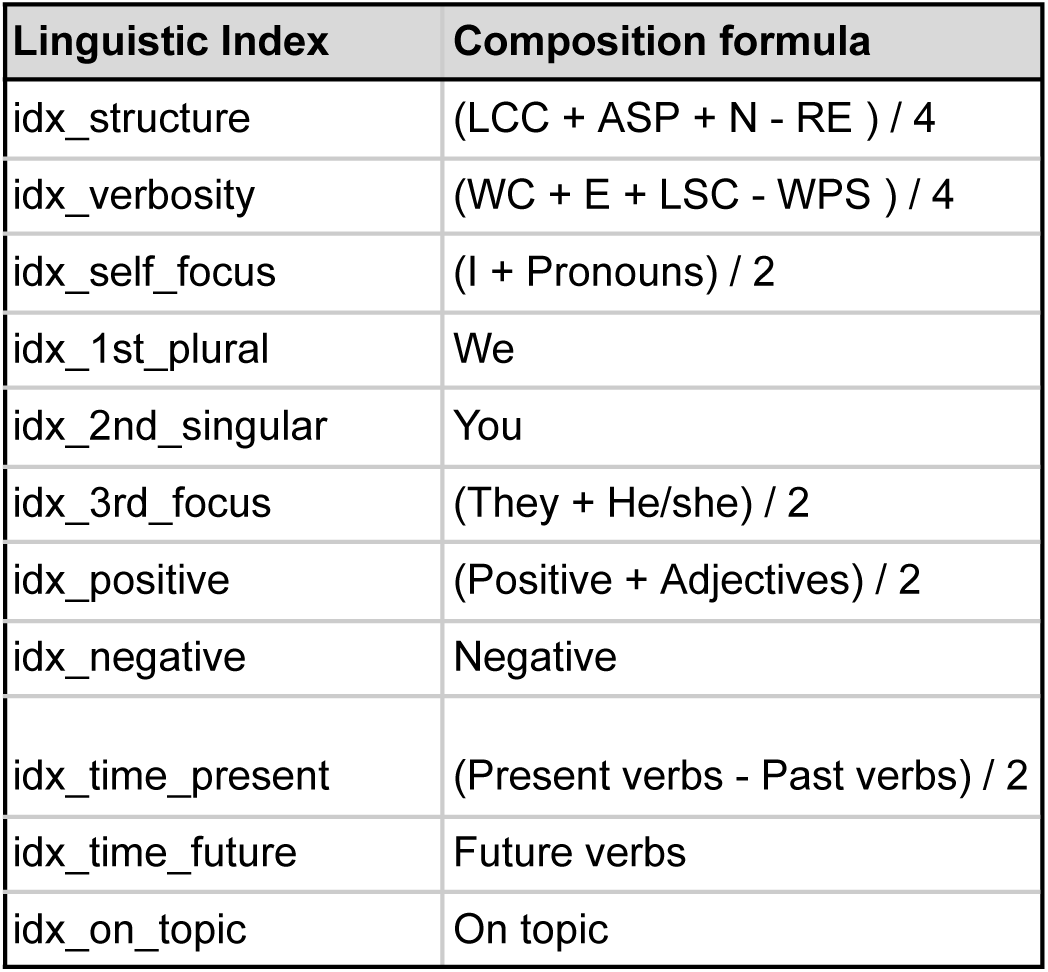
Composition formulas for the ten synthetic linguistic indices employed in the predictive model. Each index (e.g., idx_structure, idx_verbosity, idx_self_focus) was constructed through the linear combination of highly correlated structural, semantic, and morphosyntactic speech metrics identified via topological network analysis.

Using these compounded indexes along with NC and ENC variables (f1 and f2 models respectively) in a Bayesian classification framework, we tested the discrimination power of these speech markers to distinguish between risk and non-risk of symptoms and diagnosis and control groups, previously tested with the ENC index (Table 6). We reached a moderate to high specificity with a floor sensitivity of 0.8 using the f3 and f5 models for both symptom risk and diagnosis outcomes, suggesting that using different speech variables together with sociodemographic information and a more refined approach, it is possible to triage for mental risk assessment.

**Table 6.**

| Diagnostic model (case-control) |  |  |  |  |
| --- | --- | --- | --- | --- |
| Model | Prevalence (case) | AUC | Sensitivity | Specitivity |
| f1 = N + RE + LSC + ASP | 68.3% | 0.645 | 0.805 | 0.316 |
| f2 = f1 + positive + negative | 68.3% | 0.687 | 0.805 | 0.395 |
| f3 = f2 * sex + education | 68.3% | 0.800 | 0.805 | 0.632 |
| f4 = f2 * sex | 68.3% | 0.730 | 0.805 | 0.395 |
| f5 = compounded indexes * sex + education | 68.3% | 0.858 | 0.817 | 0.842 |
| Risk of psychiatric symptoms (any clinical cutoff) |  |  |  |  |
| Model | Prevalence (risk) | AUC | Sensitivity | Specitivity |
| f1 = N + RE + LSC + ASP | 57.6% | 0.608 | 0.824 | 0.140 |
| f2 = f1 + positive + negative | 57.6% | 0.675 | 0.824 | 0.440 |
| f3 = f2 * sex + education | 57.6% | 0.726 | 0.838 | 0.540 |
| f4 = f2 * sex | 57.6% | 0.677 | 0.809 | 0.480 |
| f5 = compounded indexes * sex + education | 57.6% | 0.774 | 0.809 | 0.640 |

The Bayesian logistic regression framework, optimized under a sensitivity floor of 80%, demonstrated utility as a clinical screen across all evaluated psychiatric scales (Table 7). Classification performance varied depending on the symptom and its established clinical threshold, showing moderate-to-good discriminative capacity for affective symptoms and exceptional accuracy for severe psychotic manifestations. For mood disorders, the composite linguistic indices yielded stable and balanced predictive power. In assessing anxiety severity using the GAD-7, the model achieved identical Area Under the Curve (AUC) values of 0.749 for both the mild screening threshold (gad7_5p) and the moderate threshold (gad_10p), with specificity = 0.519. Under the constrained optimization procedure, both models successfully maintained the screening requirements, yielding sensitivities of 80.3% and 80.5%, respectively. A similar screening profile was observed for depressive symptoms via the PHQ-9. For the sub-clinical threshold (phq9_8p), the model achieved an AUC of 0.801, paired with a sensitivity of 82.3% and a specificity of 55.4%. When shifting to the stricter diagnostic threshold (phq9_10p), the model’s overall discriminative capacity remained high with an AUC of 0.774. At the same time, the decision boundary adjustment significantly improved specificity to 72.9% while maintaining sensitivity at 81.2%.

**Table 7.** Performance of the Bayesian logistic classifier as a triage tool across multiple psychiatric severity thresholds. The table reports the AUC, sensitivity (optimized to maintain a strict minimum floor of ∼80%), and specificity resulting from the model’s application to predefined risk cutoffs on standardized scales for depression (PHQ-9), anxiety (GAD-7), and psychosis (PANSS subscales and total score).

| Outcome | AUC | Sensitivity | Specificity |
| --- | --- | --- | --- |
| GAD-7 5 points | 0,749 | 0,803 | 0,519 |
| GAD-7 10 points | 0,749 | 0,805 | 0,519 |
| PHQ-9 8 points | 0,801 | 0,823 | 0,554 |
| PHQ-9 10 points | 0,774 | 0,812 | 0,729 |
| PANSS positive 10 points | 0,764 | 0,818 | 0,527 |
| PANSS positive 14 points | 0,737 | 0,81 | 0,515 |
| PANSS negative 14 points | 0,776 | 0,818 | 0,552 |
| PANSS negative 20 points | 0,919 | 0,8 | 0,991 |
| PANSS total 60 points | 0,773 | 0,821 | 0,644 |
| PANSS total 75 points | 0,911 | 0,875 | 0,827 |

The predictive framework achieved its highest classification performance when distinguishing severe psychotic dimensions on the Positive and Negative Syndrome Scale (PANSS). While milder cutoff scores for positive (panss_pos_10p, AUC = 0.764; panss_pos_14p, AUC = 0.737) and negative symptoms (panss_neg_14p, AUC = 0.776) exhibited moderate discriminative profiles, with specificities hovering around 51.5%–55.2%, classification power increased in the higher-severity brackets. Specifically, for severe negative symptomatology (panss_neg_20p), the model reached an AUC of 0.919. At an optimized threshold of 0.164, it achieved a sensitivity of 80.0% with a specificity of 99.1%.

To assess the model’s discriminative capacity across different clinical thresholds, the analysis of the predicted probability distributions (Figure 4c) reveals the dynamics of group separation. At milder diagnostic cutoffs, a partial overlap between the density curves of normal and risk instances is observed, reflecting the inherent challenge of classifying subclinical symptoms. However, in higher severity brackets, most notably panss_neg_20p and panss_total_75p, the distributions exhibit greater divergence, with high specificity and AUC values above 0.91.

Furthermore, cross-validation of these risk predictions against the actual clinical diagnostic groups (Figure 4d) attests to the classifier’s validity. To further validate the group-level stratification observed in the cross-validation analysis (Figure 4d), a post hoc analysis was conducted to assess the statistical significance of the models’ predicted probabilities across distinct clinical diagnoses (Supplementary Table 2). For the affective tracking models, the classifier discriminated healthy controls from clinical cohorts. Specifically, the PHQ-9 model significantly differentiated controls from individuals with Mood Disorders (padj < 0.01), Affective Psychosis (padj < 0.001), and Non-Affective Psychosis (padj < 0.01). Similarly, the GAD-7 model effectively separated controls from both the Mood Disorders and Affective Psychosis subgroups (padj < 0.01), and detected significant differences between Affective and Non-Affective Psychosis (padj < 0.05). Conversely, models derived from the PANSS domains demonstrated targeted specificity for severe psychotic symptomatology. These models successfully discriminated healthy controls from the Affective Psychosis cohort across the Positive (padj < 0.001), Negative (padj < 0.05), and Total (padj < 0.01) symptom boundaries. This analysis shows that the composite linguistic indices not only capture generalized psychological distress but also are able to stratify distinct psychiatric subpopulations.

### 5. Stability of model performance and shift in feature importance across severity thresholds

An important finding of our predictive architecture is the decoupling of overall classification performance from individual feature dynamics. While the classifier maintains a remarkably stable discriminative capacity across a continuous spectrum of clinical cutoffs, the underlying linguistic markers driving those predictions shift dynamically with symptom severity and biological sex. As illustrated in the continuous threshold analyses, the model’s Area Under the Curve (AUC) remains resiliently bound within highly accurate tracking bands. For instance, across the entire operational range of the GAD-7 and PHQ-9, the AUC consistently hovers between 0.74 and 0.85, with no critical drop-offs even at the extreme boundaries (Supplementary Figures 1-5). This macro-level stability is similarly mirrored in psychotic symptom tracking, where the PANSS positive subscale maintains a steady baseline AUC of roughly 0.73 to 0.80, and the PANSS total curve remains bounded within a high-performance plateau of 0.75 to 0.92.

However, inspecting the internal parameter assignments reveals that the lasso-regularized feature importance is highly sensitive to symptom severity, with distinct linguistic indicators taking precedence at different stages of clinical progression (Supplementary Figures 6-10). In affective screening models, low-severity thresholds are heavily driven by temporal orientation indices. For the GAD-7 at 5 points, idx_time_present acts as the dominant positive predictor, particularly for male participants, whereas at the severe 15-point cutoff, this metric diminishes significantly, and affective load indicators like idx_negative, with positive correlation, and idx_positive, marked by an inverse, negative correlation, emerge as the primary drivers of classification. A parallel severity-dependent flip is observable in the PHQ-9 models, where conversational focus indicators, idx_on_topic and idx_2nd_singular, dominate early screening, but give way to a rise in the importance of negative emotion markers and a drop in first-person plural usage as symptoms intensify to the 15-point threshold.

This rearrangement of feature importance is even more pronounced within psychotic symptom strata, where severe symptom manifestations change the model’s linguistic tracking profile. At low-severity cutoffs (e.g., PANSS Positive at 10 points or PANSS Negative at 10 points), classification relies on localized egocentric speech metrics, marked by high positive values for idx_self_focus. However, as psychotic severity escalates to the highest thresholds of 21 points for both scales, these early markers drop out of the top rankings. For severe positive symptoms (PANSS+ 21 points), the model pivots toward conversational markers, shifting idx_2nd_singular to a positive predictor and idx_3rd_focus to a negative predictor. For severe negative symptoms (PANSS-21 points), idx_negative increases exponentially, dominating the model’s feature space and registering a scaled importance near 0.75. Collectively, these results demonstrate that while the machine learning framework serves as a reliable, stable triage instrument across all continuous cutoffs, keeping its AUC above 0.7 in any scale cutoff with representativity above 8% in the study population, and it does so by mathematically adapting to a shifting linguistic phenotyping architecture that evolves with clinical severity.

## DISCUSSION

The present study introduces a comprehensive framework for assessing cognitive development, socioemotional functioning, and mental health through the analysis of spontaneous speech. By combining graph-theoretical measures of discourse organization with psycholinguistic and emotional features, we demonstrate that language can be quantified along developmental trajectories that reflect expected educational maturation and subsequently leveraged for mental health screening and symptom-informed clinical assessment. Rather than proposing isolated language markers, our findings support a hierarchical pipeline in which language serves as a digital marker across the continuum from developmental monitoring to preventive screening and clinical decision support.

Our first objective was to establish normative developmental references for language organization across formal education. Both Narrative Complexity (NC) and Emotional Narrative Complexity (ENC) exhibited nonlinear growth that was accurately described by asymptotic exponential functions. This replicates in large scale what was demonstrated in Mota et al. (2018) in a population of 135 typical participants. Moreover, it demonstrates that emotional content also develops with educational attainment, a correlation also described in a first-episode psychosis population (Mota, Ribeiro, et al., 2023). This finding supports the hypothesis that language organization develops through rapid gains during early schooling followed by gradual stabilization, consistent with general models of cognitive development and educational learning.

The estimated characteristic times (τ = 7.77 years for NC and τ = 6.97 years for ENC) indicate that narrative abilities approach their mature organization during the final years of Brazilian elementary education. The asymptotic behavior suggests that schooling progressively refines linguistic organization until reaching a relatively stable level, reflecting the cumulative acquisition of literacy, syntactic complexity, discourse planning, and abstract reasoning. Importantly, these parameters should not be interpreted as developmental endpoints, but rather as indicators of the educational pace through which increasingly sophisticated narrative structures emerge.

These findings reinforce the view that language organization is not solely determined by biological maturation. Instead, formal education appears to play a central role in shaping the cognitive mechanisms required for producing coherent, hierarchically organized narratives. Exposure to written language, increasingly complex texts, classroom discourse, and metacognitive activities likely contributes to the progressive refinement captured by NC and ENC.

Although both indices followed similar developmental trajectories, they represent distinct aspects of language production.

Narrative Complexity primarily captures the structural organization of discourse. By integrating graph attributes related to lexical diversity, graph size, and long- and short-range recurrences, NC reflects the ability to organize ideas coherently over the course of a narrative. Rather than measuring vocabulary size or verbal fluency alone, the index quantifies an emergent property of discourse organization that reflects planning, working memory, and logical integration of information (Mota et al., 2016, 2018, 2020; Mota, Weissheimer, et al., 2023).

Emotional Narrative Complexity extends this structural representation by incorporating emotional valence information. These results replicate in a Brazilian large-scale population the association between emotional expression and schooling already mapped in other cultural contexts (Yuan et al., 2024). Consequently, ENC captures the degree to which affective information is coherently integrated into narrative production. This additional emotional dimension likely reflects higher-order socioemotional processes, including emotional regulation, autobiographical integration, and social cognition.

A major contribution of this work is the establishment of percentile-based normative curves calibrated by educational attainment. Because language performance is strongly influenced by schooling, interpreting raw speech measures without accounting for educational level may lead to substantial diagnostic bias. Lower narrative complexity in individuals with limited educational exposure may simply reflect expected developmental variation rather than pathology, whereas highly educated individuals may compensate for subtle cognitive impairment through preserved linguistic abilities.

The normative curves proposed here address this limitation by providing developmental expectations analogous to pediatric growth charts. Just as anthropometric curves allow clinicians to determine whether physical growth follows expected developmental trajectories, NC and ENC percentiles enable the assessment of whether language organization and socioemotional narrative development are consistent with an individual’s educational history.

This normative approach substantially increases the interpretability and potential scalability of language-based biomarkers. Instead of relying on absolute values, clinicians and researchers can interpret each individual’s performance relative to an expected developmental distribution, facilitating early identification of atypical developmental trajectories.

ENC fails to discriminate between individuals with preserved mental wellness from those presenting clinical symptoms. Although the observed AUC values were low, the consistently high sensitivity suggests that ENC is particularly suitable as a first-line screening tool for mental wellness, identifying atypical developmental trajectories.

From a public health perspective, screening instruments prioritize sensitivity over specificity to minimize missed cases requiring further evaluation. The relatively modest specificity observed in the present study indicates that ENC should not be interpreted as a standalone diagnostic instrument. Rather, it serves as an efficient population-level radar system that identifies individuals who may benefit from a more comprehensive clinical assessment.

This interpretation aligns with the conceptual purpose of the framework. Rather than replacing standardized psychiatric evaluation, ENC provides an objective, scalable, and low-burden method for identifying individuals whose emotional narrative organization deviates from expected developmental patterns. Such an approach may prove particularly valuable in educational systems, primary care, and occupational health settings, where rapid large-scale monitoring is often necessary, but specialist availability remains limited.

The second major contribution of this study is the demonstration that spontaneous speech contains clinically meaningful information that extends beyond general screening. Machine-learning models that combine graph-derived structural measures with psycholinguistic and emotional features achieved AUC values ranging from 0.74 to 0.92 across multiple psychometric scales and symptom-severity thresholds, while maintaining high sensitivity.

Importantly, these models estimate symptom dimensions rather than categorical diagnoses. This distinction is consistent with contemporary dimensional approaches to psychopathology, in which psychiatric symptoms exist along continuous spectra rather than discrete diagnostic boundaries (Conway et al., 2022; Cuthbert & Insel, 2013; Kotov et al., 2017). Depressive and anxiety syndromes appear to share overlapping alterations in speech content, broadly characterized by reduced verbal informativeness and shorter words. Depressive syndromes may additionally be associated with more frequent punctuation, more straightforward sentence constructions, reduced linguistic complexity, and potentially longer or more frequent pauses. In contrast, psychotic syndromes appear to affect both the structure and content of discourse more substantially. These alterations include reduced lexical richness and linguistic complexity, lower verbal informativeness, shorter sentences, more frequent punctuation—suggesting simpler syntactic structures and potentially more frequent pauses—and a greater use of the present tense.

Negative psychotic symptoms further reproduce the marked reduction in the connectivity and size of word recurrence graphs described by Mota, Weissheimer, et al. (2023) alongside a lower frequency of second-person pronouns, suggesting reduced linguistic orientation toward the interlocutor. Conversely, positive and general psychotic symptoms are associated with greater self-referentiality, reflected in more frequent use of first-person pronouns, as well as shorter words.

Taken together, these findings suggest that speech patterns may reflect the phenomenological similarities between depressive and anxiety syndromes, while psychotic syndromes are associated with more pronounced alterations in discourse organization and communicative structure. This distinction highlights the potential of linguistic features to capture both shared and syndrome-specific dimensions of psychopathology.

Furthermore, the observation that individual linguistic variables differed across symptom-severity levels suggests that distinct speech features may capture partially dissociable dimensions of psychopathology. Previous computational studies have shown that linguistic characteristics such as syntactic complexity, semantic coherence, referential cohesion, lexical choices, and pronoun use are associated with clinically relevant features of psychiatric disorders, particularly psychosis (Corcoran et al., 2020; Low et al., 2020; Morgan et al., 2021). These findings support the interpretation of structural organization, emotional content, temporal references, and self-referential language as complementary linguistic dimensions that may reflect different aspects of psychological functioning, although their correspondence to partially independent neurocognitive mechanisms remains to be established. Such interpretability represents a potential advantage of feature-based approaches over less interpretable black-box models, as it allows computational findings to be related to phenomenological constructs routinely assessed during psychiatric interviews (Malgaroli et al., 2023; Viduani et al., 2026).

Taken together, these findings support a unified framework that integrates developmental monitoring, preventive screening, and clinical assessment through spontaneous speech analysis.

Several limitations should be acknowledged. First, the normative curves were derived from cross-sectional rather than longitudinal data. Although the large sample provides robust estimates of developmental trajectories, longitudinal studies will be necessary to characterize individual developmental dynamics more precisely. Second, educational attainment was used as the primary developmental reference. While schooling represents one of the strongest determinants of language organization, additional variables—including literacy quality, socioeconomic status, bilingualism, and cultural background—may further influence narrative development and should be incorporated into future normative models.

Third, although the clinical sample included multiple psychiatric diagnoses, its size remains relatively modest for developing highly specialized diagnostic models. Independent multicenter validation involving larger and more diverse clinical populations will be essential to establish the generalizability of symptom prediction across healthcare settings. Finally, the moderate specificity observed for ENC indicates that the index should be interpreted as a screening measure, with its greatest value when integrated with complementary clinical assessments.

Future work should focus on longitudinal monitoring of individuals across educational and clinical settings, external validation in diverse linguistic populations, and integration with digital health infrastructures. Because the proposed framework relies on brief speech samples collected through low-cost protocols, it offers considerable potential for implementation in schools, primary healthcare services, telemedicine platforms, and population-based mental health surveillance programs.

This study demonstrates that spontaneous speech contains sufficient information to characterize normative cognitive and socioemotional development, identify deviations from typical trajectories associated with mental wellness, and estimate clinically relevant symptom dimensions. The establishment of percentile-based developmental curves for Narrative Complexity and Emotional Narrative Complexity provides interpretable normative references analogous to growth charts for language development. When combined with machine-learning models calibrated to standardized psychiatric scales, these language-based biomarkers constitute a unified framework spanning developmental monitoring, preventive mental health screening, and symptom-informed clinical decision support. Collectively, these findings establish spontaneous speech as a scalable, interpretable, and clinically meaningful digital biomarker with broad applications in education, public health, and mental healthcare.

## Data Availability

The datasets generated and analyzed in this study are not publicly available due to ethical and privacy restrictions concerning human participant data. De-identified data supporting the findings of this study may be made available to qualified researchers upon reasonable request to the corresponding author, subject to approval by the relevant ethics committees and compliance with applicable data protection regulations and institutional agreements.

---

**Supplementary Figure 1.**
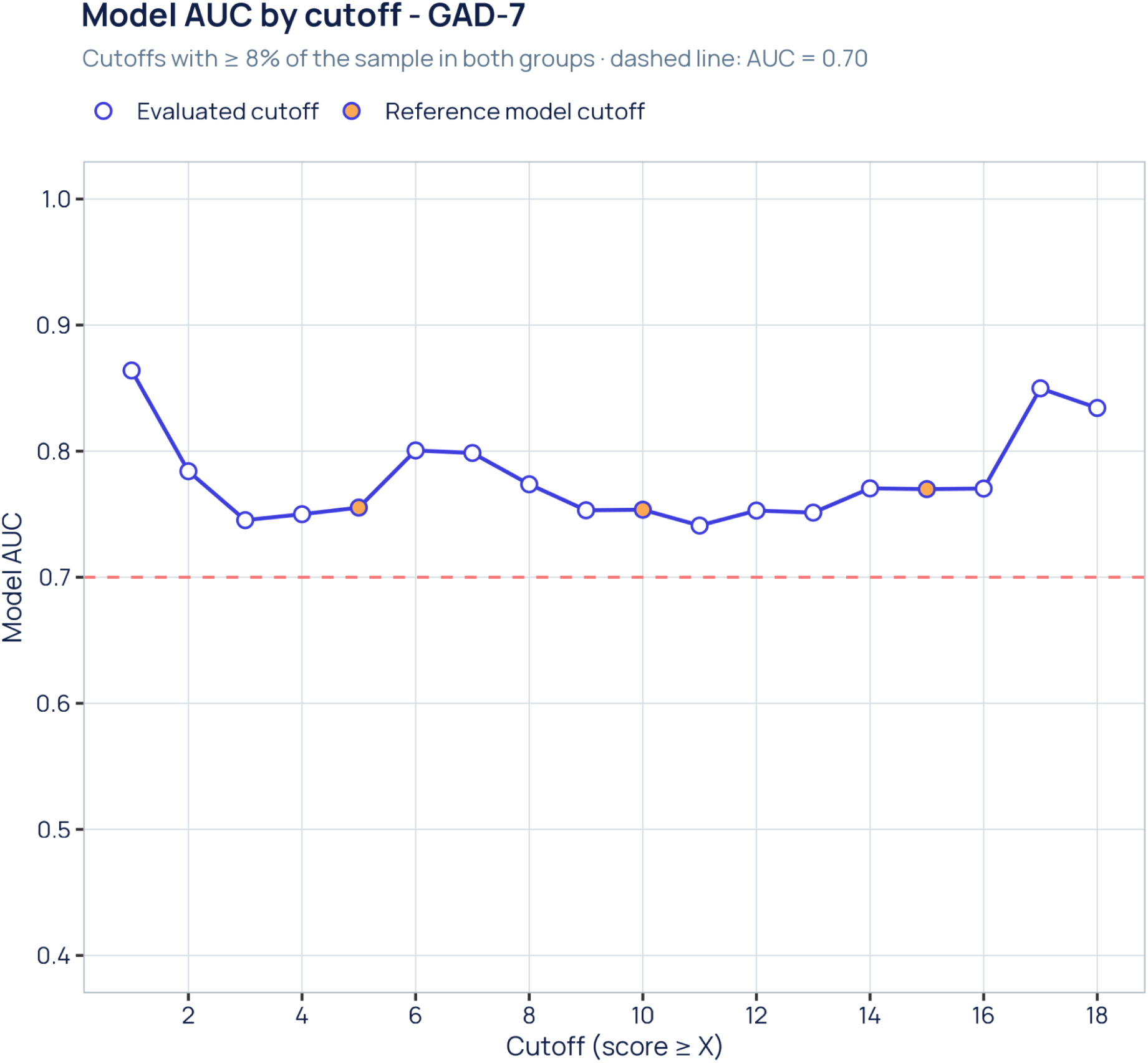
Model discrimination across GAD-7 cutoffs. Area under the ROC curve (AUC) of the lasso-regularized Bayesian logistic model trained to classify participants at or above each GAD-7 cutoff (score ≥ X), evaluated in steps of one point. Only cutoffs leaving at least 8% of the sample (n = 118) in both the at-risk and reference groups are shown (cutoffs 1–18). Orange points mark the cutoffs of the reference models shown in the feature-importance figure (5, 10 and 15 points). The dashed line marks AUC = 0.70. AUC stayed between 0.74 and 0.86 at every cutoff, with its lowest values in the mid-range (minimum at ≥ 11). AUCs are in-sample (apparent) estimates.

**Supplementary Figure 2.**
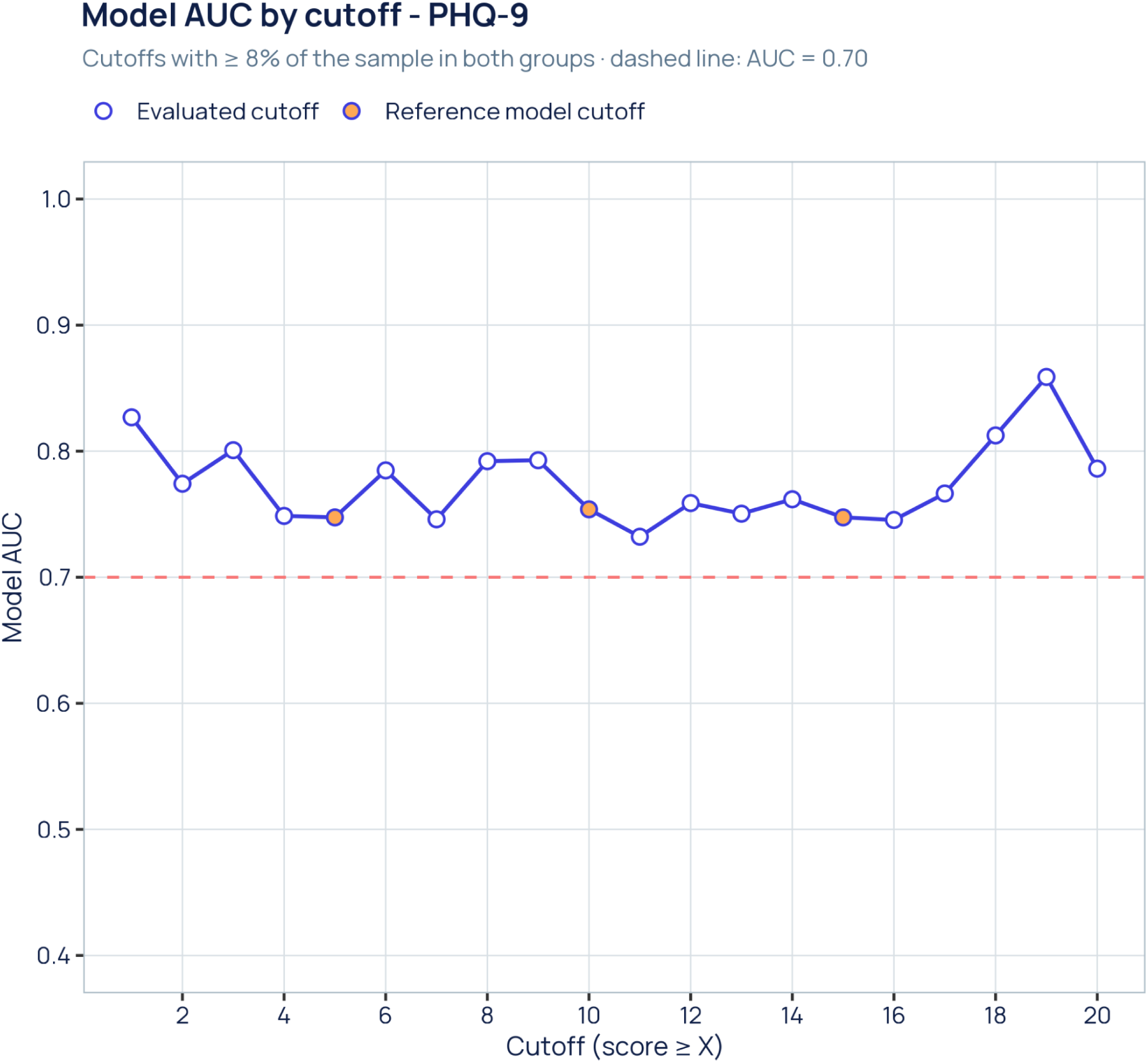
Model discrimination across PHQ-9 cutoffs. Same analysis as Supplementary Figure 1, applied to the PHQ-9 (cutoffs 1–20). Orange points mark the reference cutoffs at 5, 10 and 15 points (AUC = 0.75, 0.75 and 0.75). The dashed line marks AUC = 0.70. AUC ranged from 0.73 (≥ 11) to 0.86 (≥ 19) and never fell below the 0.70 line. AUCs are in-sample (apparent) estimates.

**Supplementary Figure 3.**
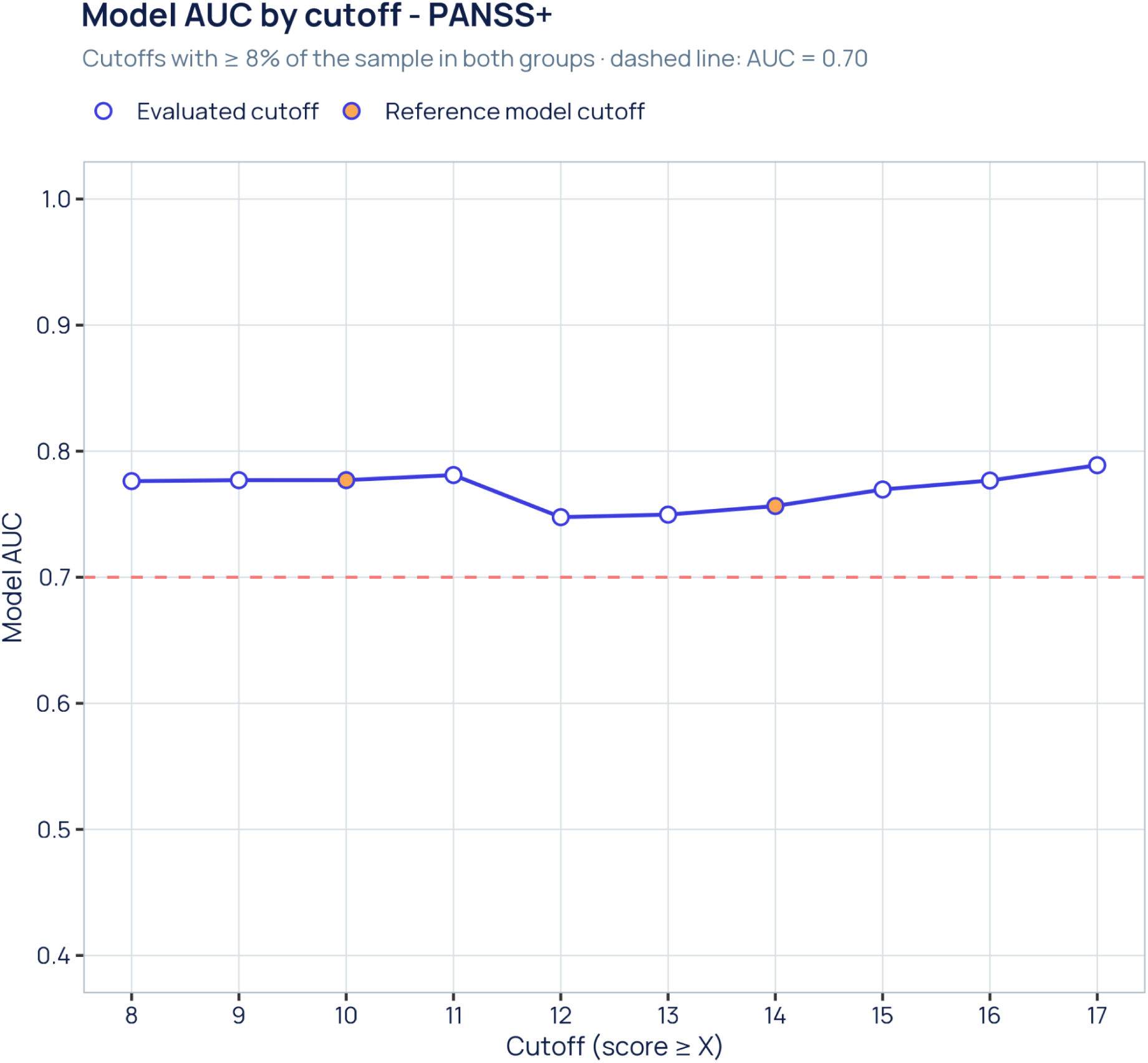
Model discrimination across PANSS positive subscale cutoffs. Same analysis as Supplementary Figure 1, applied to the PANSS positive subscale (PANSS+). Only cutoffs 8–17 met the 8% representation criterion, so the 21-point reference model is outside the evaluable range. Orange points mark the 10- and 14-point reference cutoffs. The dashed line marks AUC = 0.70. Performance was the flattest of all scales, with AUC between 0.75 and 0.79. AUCs are in-sample (apparent) estimates.

**Supplementary Figure 4.**
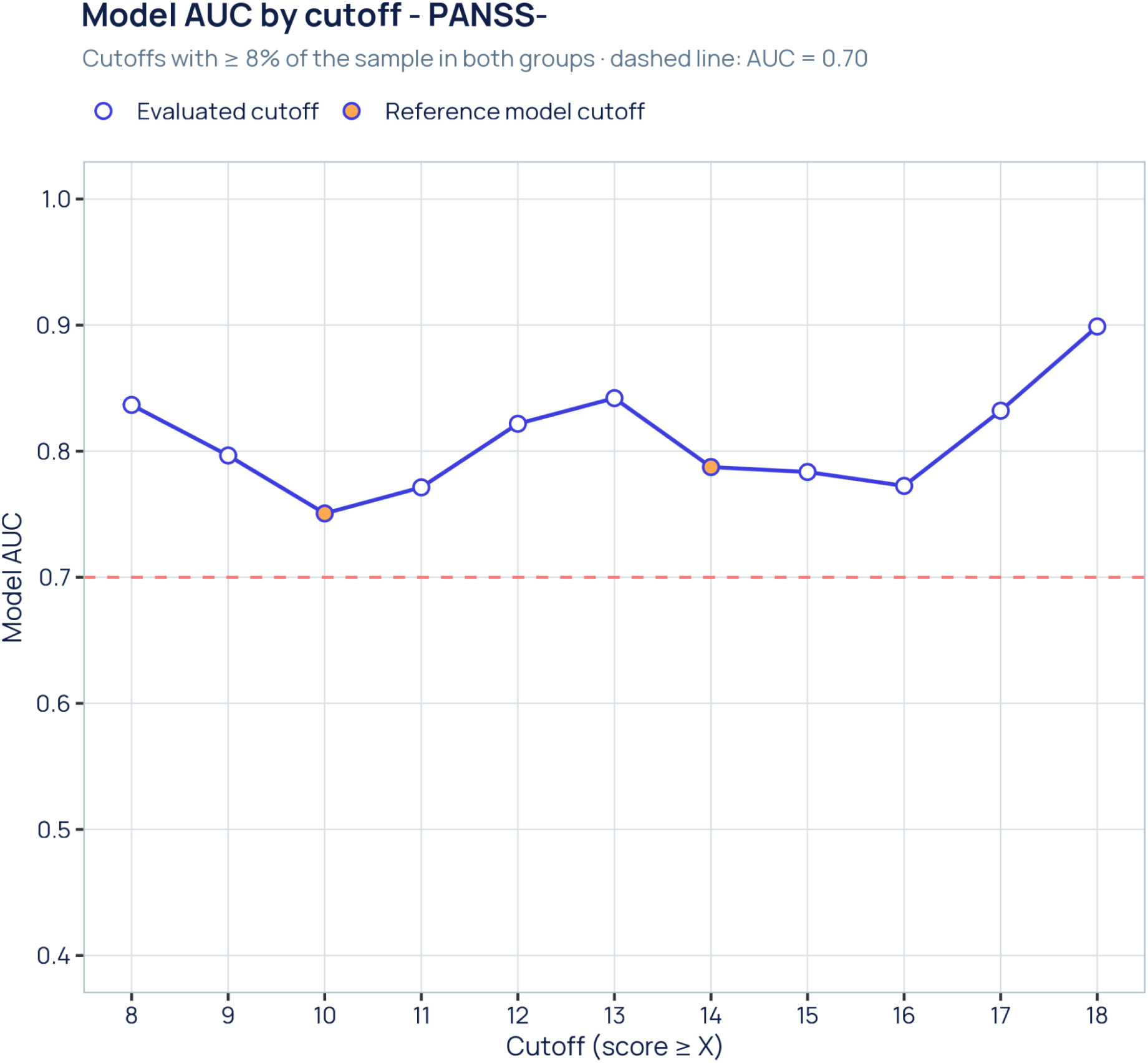
Model discrimination across PANSS negative subscale cutoffs. Same analysis as Supplementary Figure 1, applied to the PANSS negative subscale (PANSS-), cutoffs 8–18. Orange points mark the 10- and 14-point reference cutoffs (AUC = 0.75 and 0.79); the 21-point cutoff did not meet the 8% criterion. The dashed line marks AUC = 0.70. AUC ranged from 0.75 (≥ 10) to 0.90 (≥ 18). AUCs are in-sample (apparent) estimates.

**Supplementary Figure 5.**
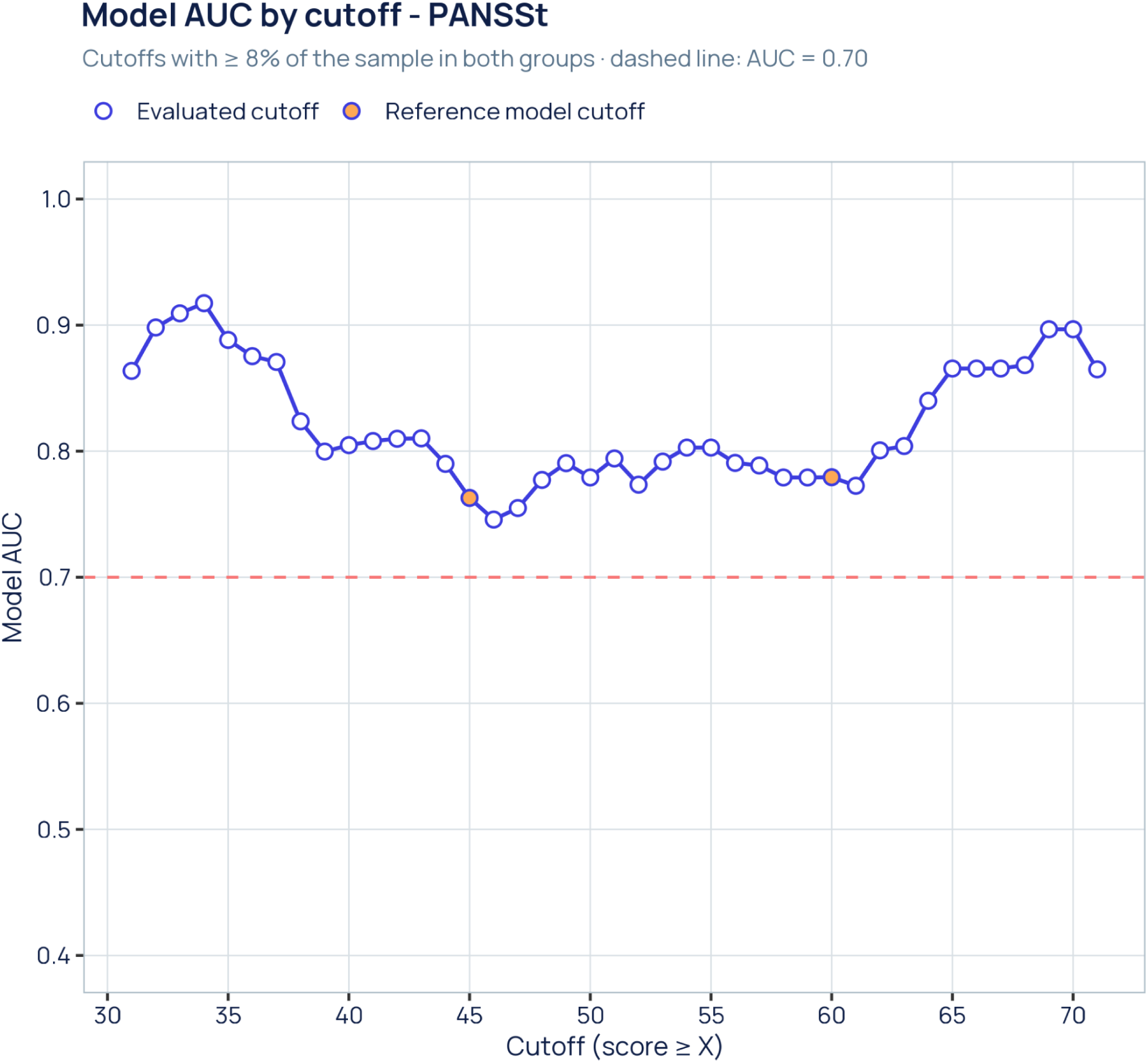
Model discrimination across PANSS total score cutoffs. Same analysis as Supplementary Figure 1, applied to the PANSS total score (PANSSt), cutoffs 31–71. Orange points mark the 45- and 60-point reference cutoffs (AUC = 0.76 and 0.78); the 75-point cutoff did not meet the 8% criterion. The dashed line marks AUC = 0.70. AUC followed a U-shaped profile from 0.75 to 0.92: highest at the extremes of the range (peak at ≥ 34) and lowest at mid-range cutoffs (minimum at ≥ 46). AUCs are in-sample (apparent) estimates.

**Supplementary Figure 6.**
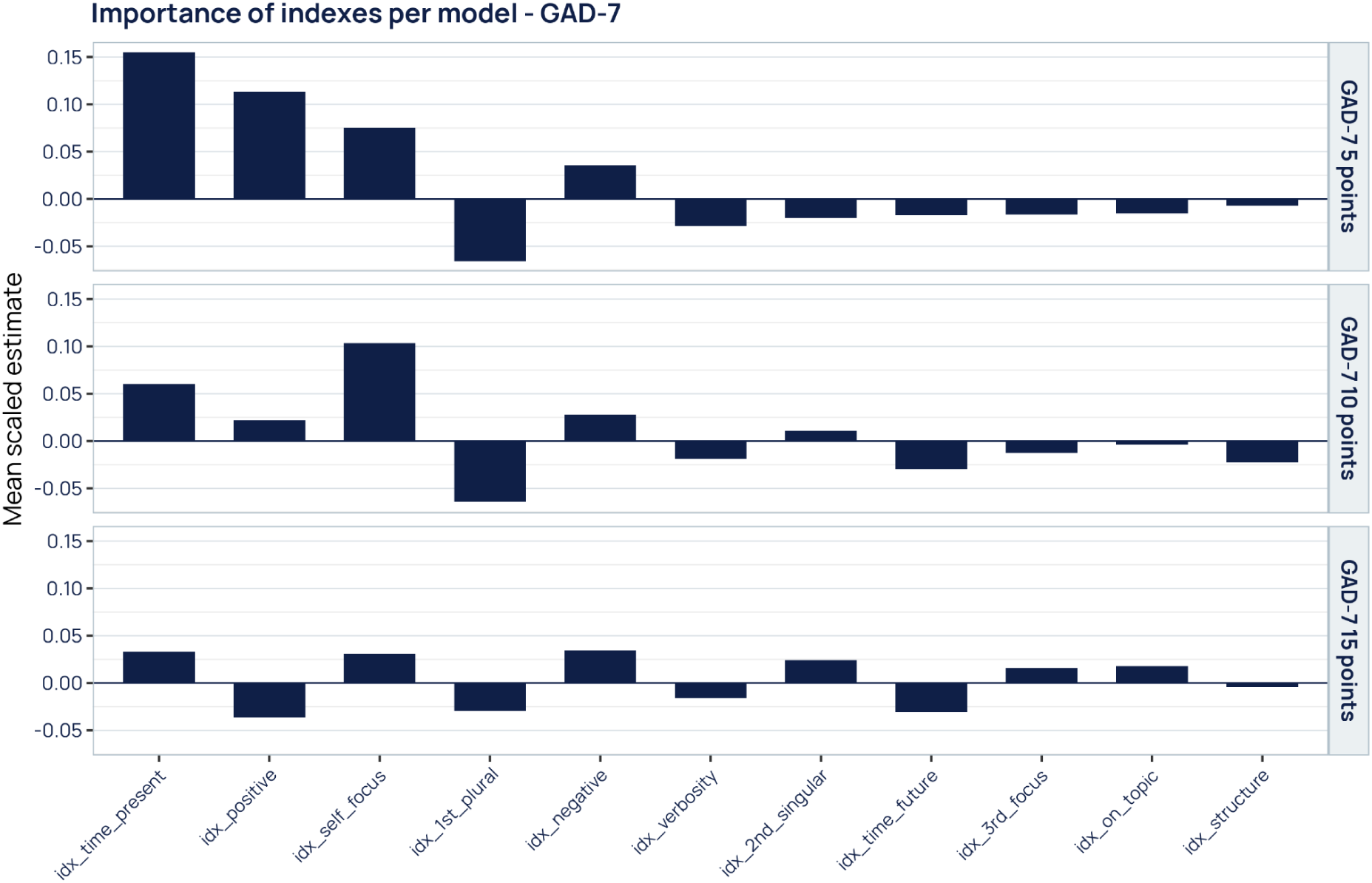
Linguistic index weights across GAD-7 severity cutoffs. Posterior mean coefficients of the lasso-regularized Bayesian logistic models classifying participants at or above 5, 10 and 15 GAD-7 points (rows). Indices were standardized before fitting, so bar heights are comparable within and across panels (log-odds per SD). Each bar is the average of the female and male slopes (main effect and main effect + sex interaction). Indices are ordered by absolute importance in the 5-point model. At the mildest cutoff, present-oriented temporal focus (+0.16) and positive affect (+0.11) dominate. At 10 points, self-focus becomes the strongest marker (+0.10), alongside fewer first-person plural references (−0.06). At 15 points, weights flatten, and positive affect reverses sign (−0.04) while negative affect remains positive (+0.04).

**Supplementary Figure 7.**
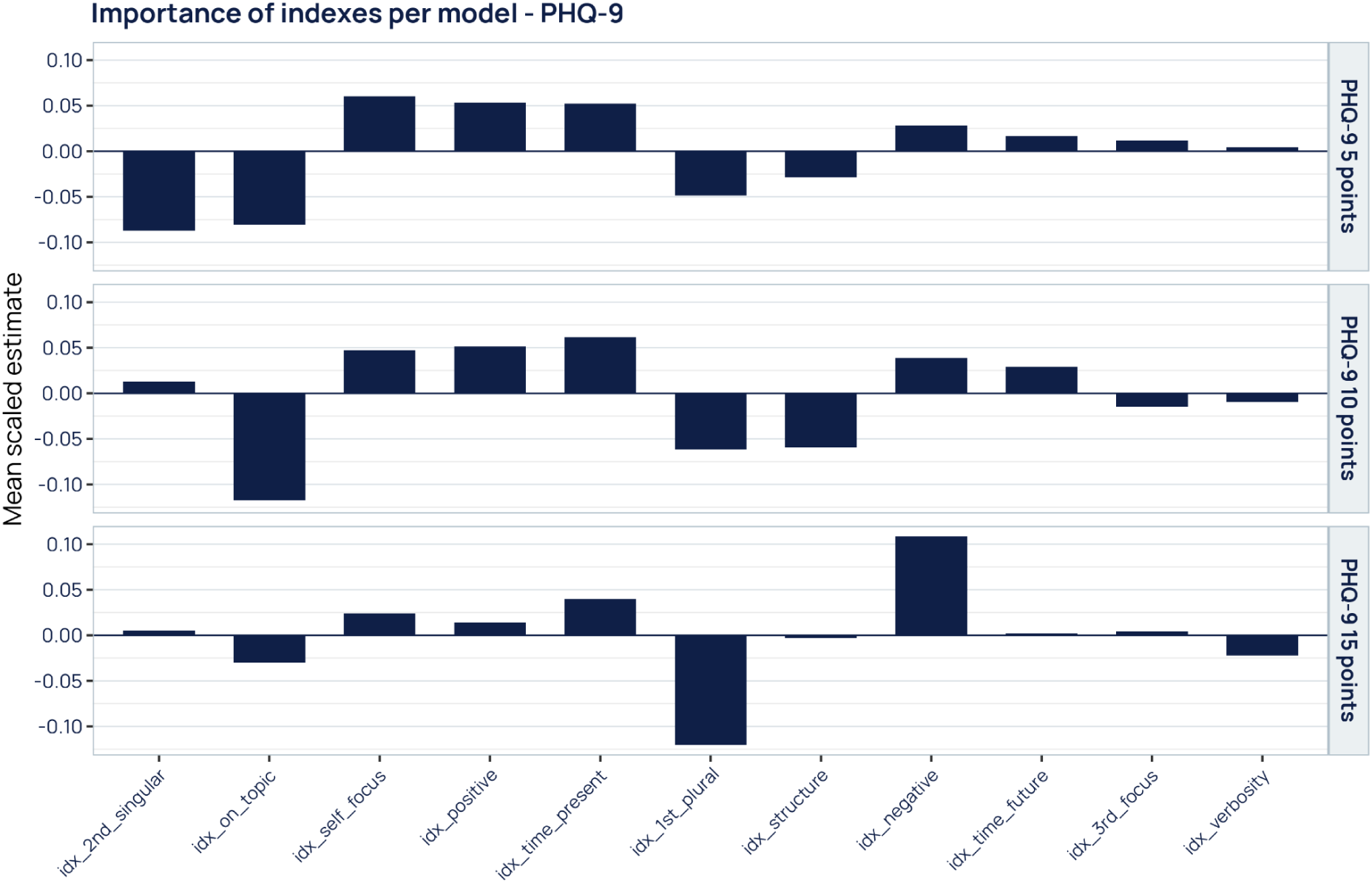
Linguistic index weights across PHQ-9 severity cutoffs. Same as Supplementary Figure 6, for models at 5, 10 and 15 PHQ-9 points. Indices are ordered by absolute importance in the 5-point model. At the mildest cutoff, fewer second-person singular references (−0.09) and lower topic adherence (−0.08) carry the most weight. At 10 points, lower topic adherence is the leading marker (−0.12). At 15 points, the profile shifts to fewer first-person plural references (−0.12) and more negative affect (+0.11).

**Supplementary Figure 8.**
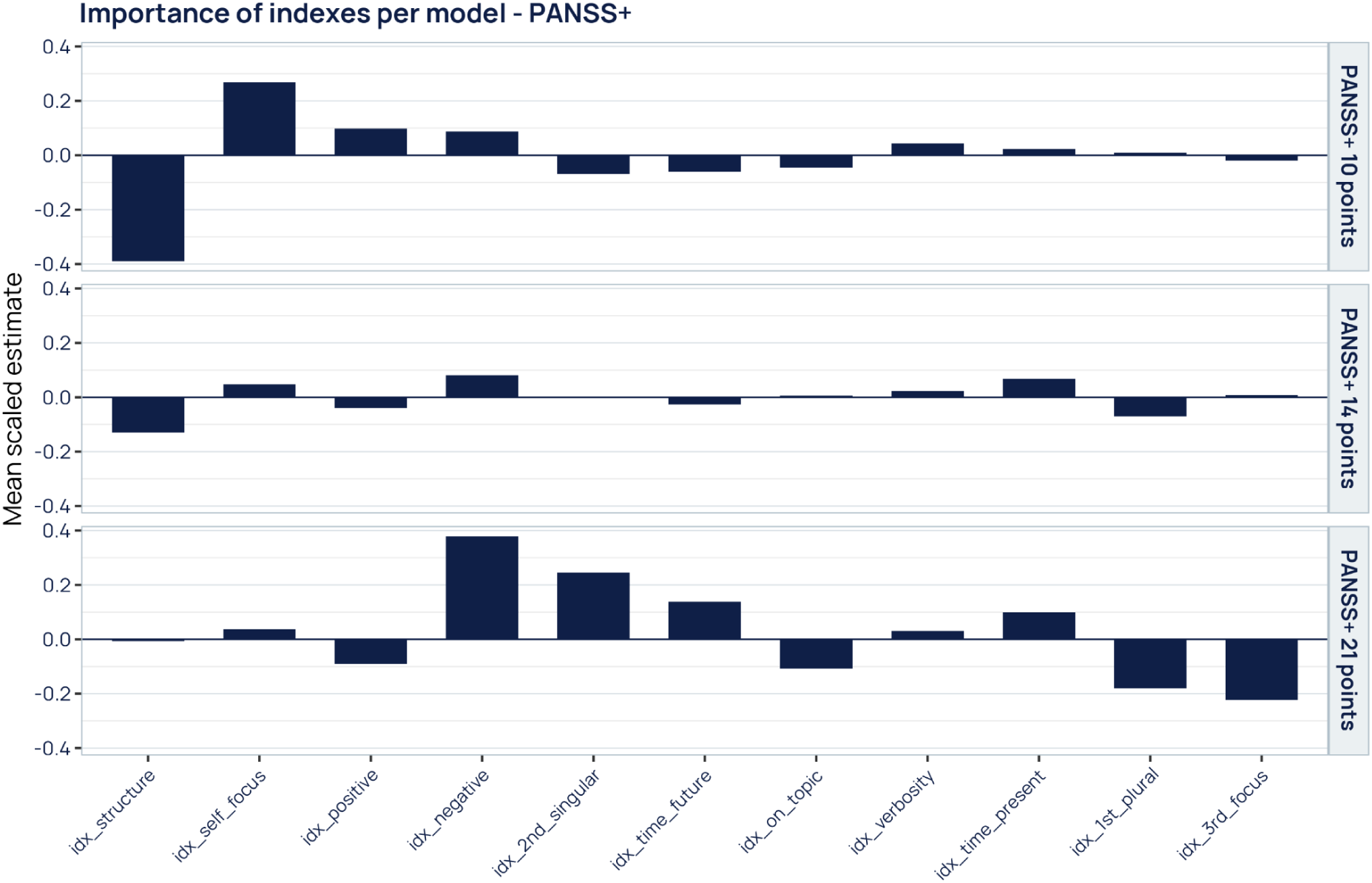
Linguistic index weights across PANSS positive subscale cutoffs. Same as Supplementary Figure 6, for models at 10, 14 and 21 PANSS+ points. Indices are ordered by absolute importance in the 10-point model. At 10 points, lower discourse structure (−0.39) and higher self-focus (+0.27) dominate. Both weaken at 14 points (−0.13 and +0.05). At 21 points, the model relies instead on negative affect (+0.38), second-person singular references (+0.25) and fewer third-person references (−0.22).

**Supplementary Figure 9.**
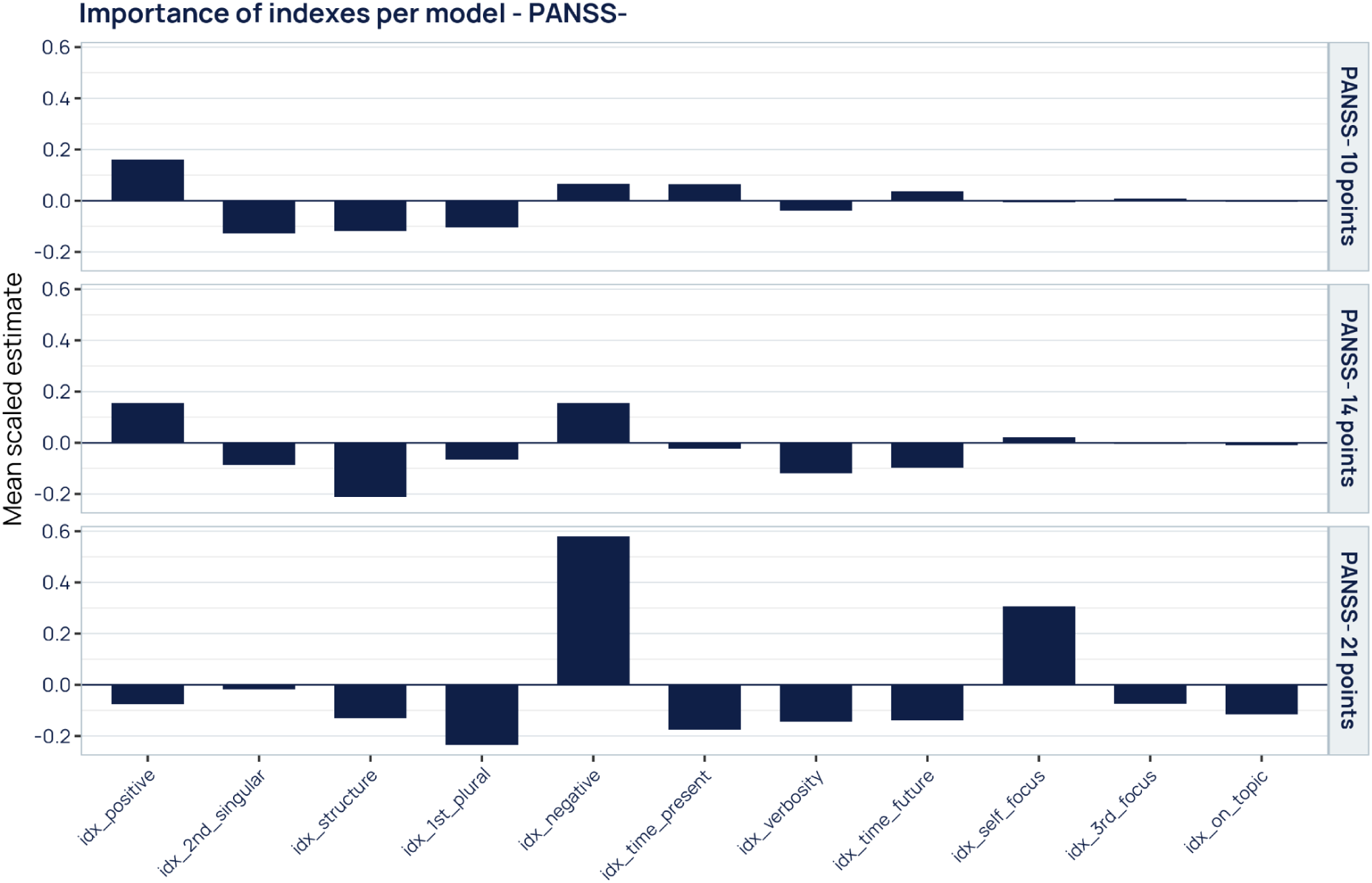
Linguistic index weights across PANSS negative subscale cutoffs. Same as Supplementary Figure 6, for models at 10, 14 and 21 PANSS-points. Indices are ordered by absolute importance in the 10-point model. At 10 points, the main markers are positive affect (+0.16), fewer second-person singular references (−0.13) and lower discourse structure (−0.12). At 14 points, lower structure becomes the leading marker (−0.21). At 21 points, negative affect (+0.58) and self-focus (+0.31) carry the largest weights in any model, together with fewer first-person plural references (−0.24).

**Supplementary Figure 10.**
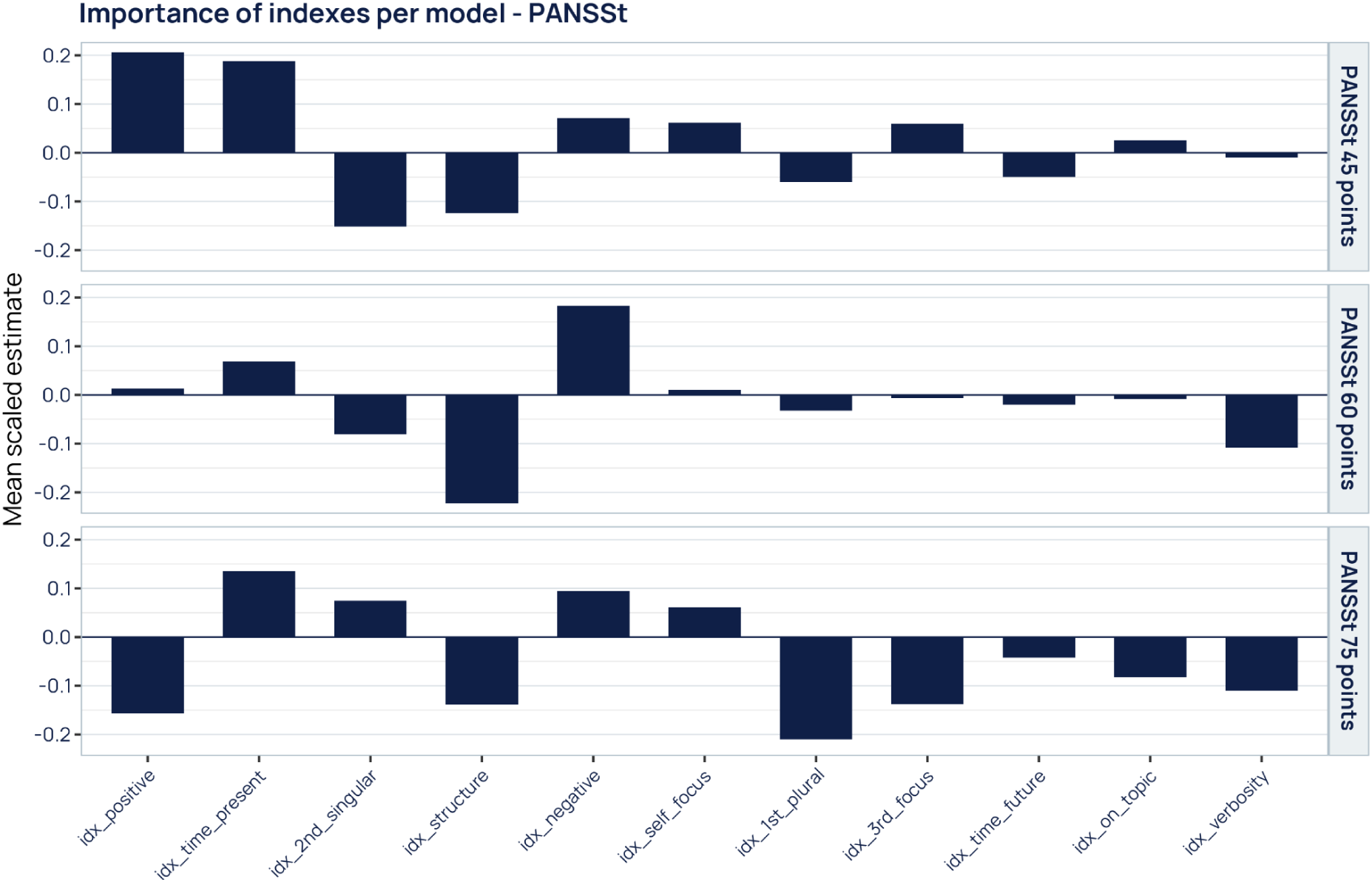
Linguistic index weights across PANSS total score cutoffs. Same as Supplementary Figure 6, for models at 45, 60 and 75 PANSSt points. Indices are ordered by absolute importance in the 45-point model. At 45 points, the model is driven by positive affect (+0.21) and present-oriented temporal focus (+0.19). At 60 points, lower discourse structure (−0.22) and negative affect (+0.18) lead. At 75 points, positive affect reverses sign (−0.16), and fewer first-person plural references become the strongest marker (−0.21).

**Supplementary Table 1.**
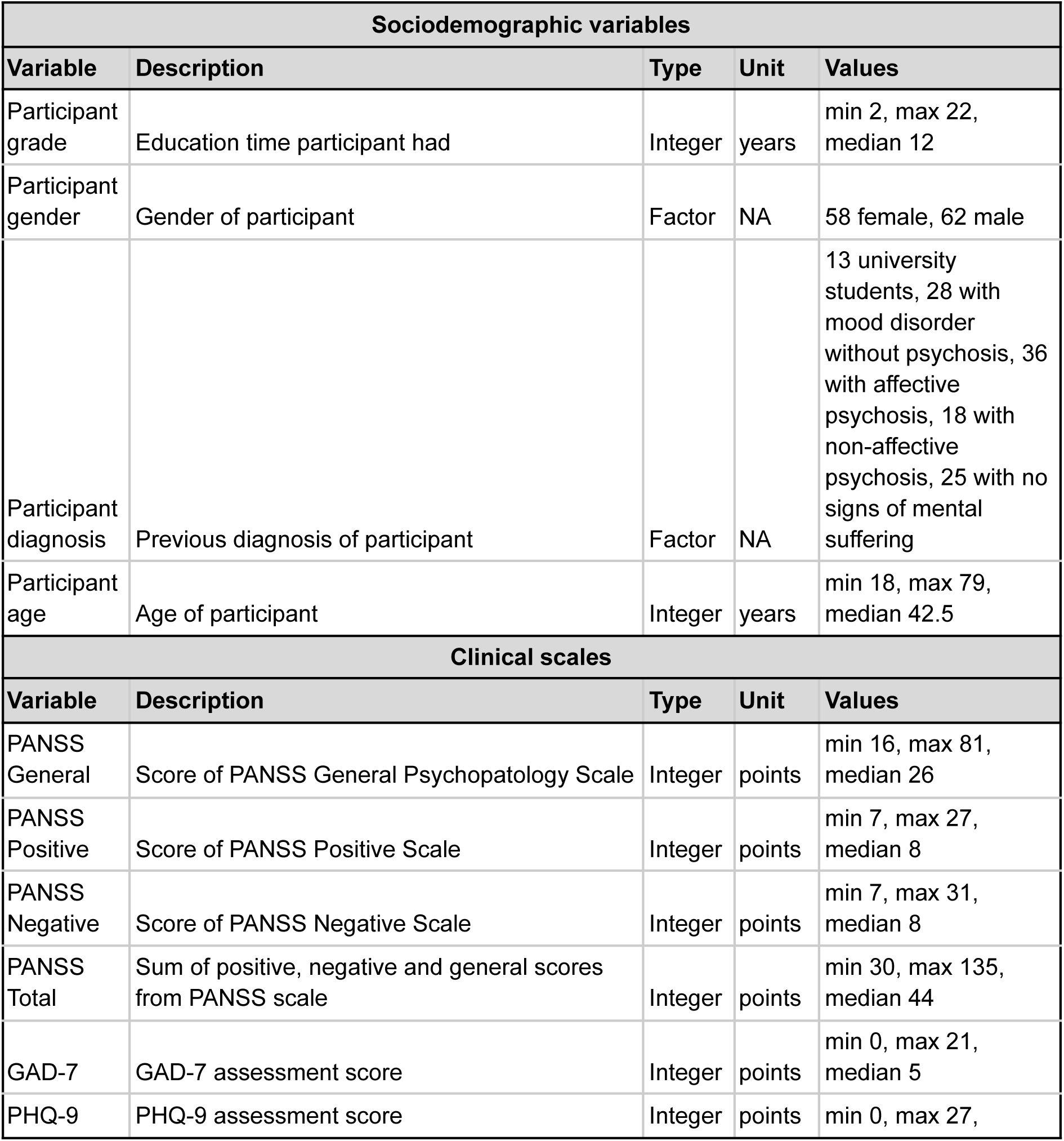
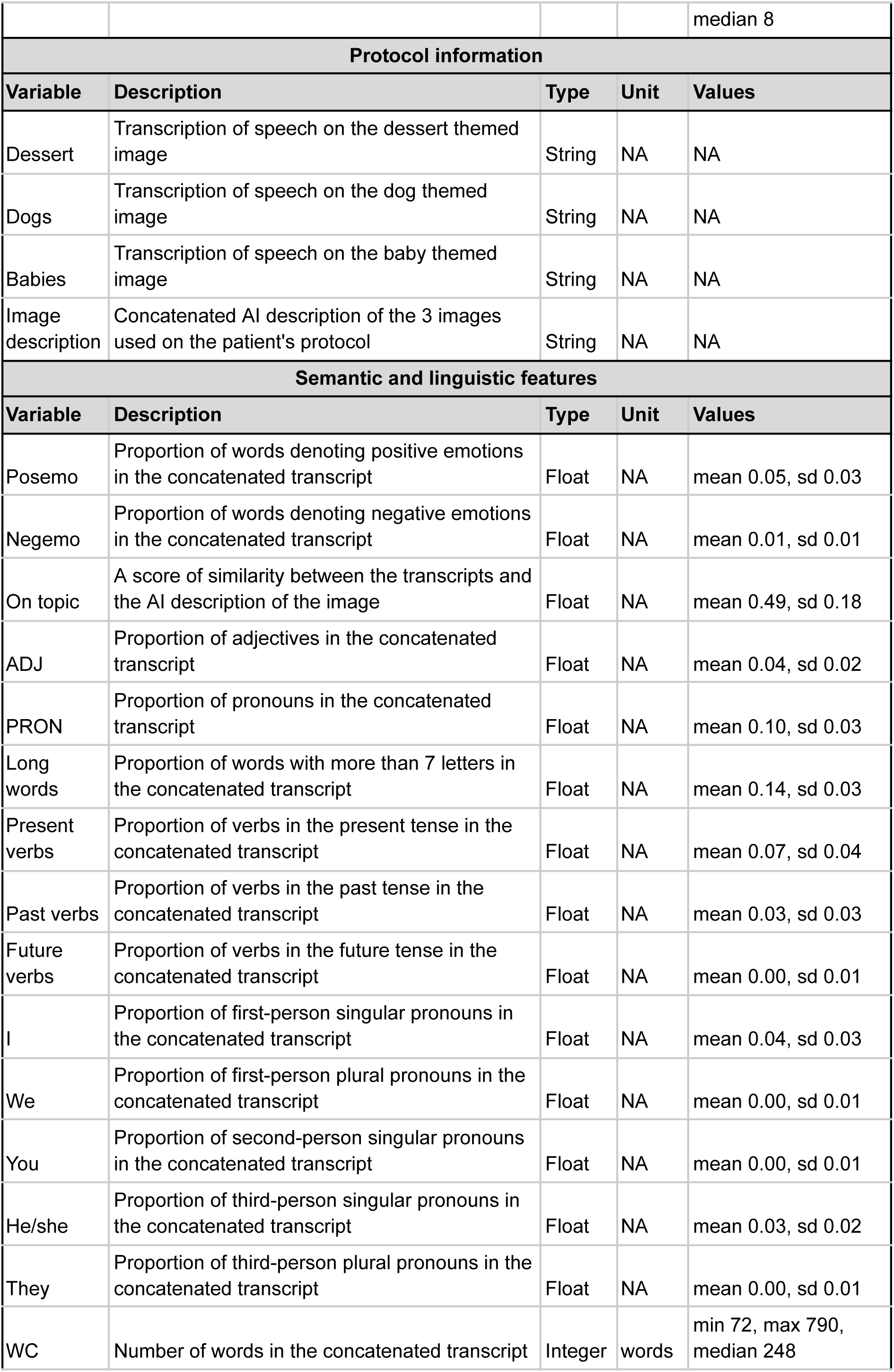
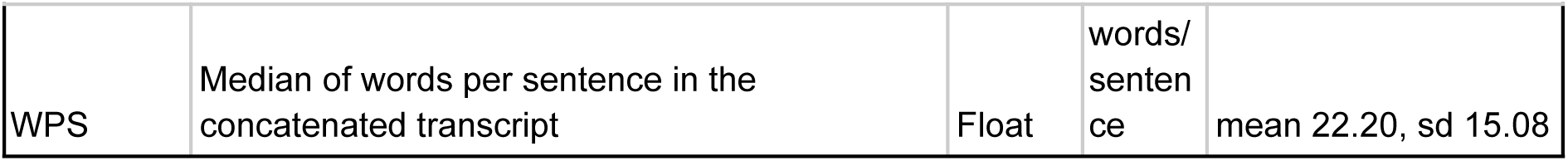
Sociodemographic, clinical and linguistic characteristics of the sample. Variables are grouped into sociodemographic data, clinical scale scores, protocol material and semantic/linguistic features extracted from the speech transcripts. For each variable, the table gives its description, data type, unit and summary statistics: median and range for integer variables, mean and standard deviation for proportions, and counts for categorical variables. Speech was elicited by asking participants to describe three images (dessert, dogs and babies). The three transcripts were concatenated per participant before feature extraction. Lexical and morphosyntactic features are proportions of the total word count of the concatenated transcript. “On topic” is the semantic similarity between each participant’s transcript and an AI-generated description of the same images. PANSS, Positive and Negative Syndrome Scale; GAD-7, Generalized Anxiety Disorder 7-item scale; PHQ-9, Patient Health Questionnaire 9-item; WC, word count; WPS, words per sentence; ADJ, adjectives; PRON, pronouns.

| Sociodemographic variables |  |  |  |  |
| --- | --- | --- | --- | --- |
| Variable | Description | Type | Unit | Values |
| Participant grade | Education time participant had | Integer | years | min 2, max 22, median 12 |
| Participant gender | Gender of participant | Factor | NA | 58 female, 62 male |
| Participant diagnosis | Previous diagnosis of participant | Factor | NA | 13 university students, 28 with mood disorder without psychosis, 36 with affective psychosis, 18 with non-affective psychosis, 25 with no signs of mental suffering |
| Participant age | Age of participant | Integer | years | min 18, max 79, median 42.5 |
| Clinical scales |  |  |  |  |
| Variable | Description | Type | Unit | Values |
| PANSS General | Score of PANSS General Psychopathology Scale | Integer | points | min 16, max 81, median 26 |
| PANSS Positive | Score of PANSS Positive Scale | Integer | points | min 7, max 27, median 8 |
| PANSS Negative | Score of PANSS Negative Scale | Integer | points | min 7, max 31, median 8 |
| PANSS Total | Sum of positive, negative and general scores from PANSS scale | Integer | points | min 30, max 135, median 44 |
| GAD-7 | GAD-7 assessment score | Integer | points | min 0, max 21, median 5 |
| PHQ-9 | PHQ-9 assessment score | Integer | points | min 0, max 27, |

|  |  |  |  | median 8 |
| --- | --- | --- | --- | --- |
| Protocol information |  |  |  |  |
| Variable | Description | Type | Unit | Values |
| Dessert | Transcription of speech on the dessert themed image | String | NA | NA |
| Dogs | Transcription of speech on the dog themed image | String | NA | NA |
| Babies | Transcription of speech on the baby themed image | String | NA | NA |
| Image description | Concatenated AI description of the 3 images used on the patient's protocol | String | NA | NA |
| Semantic and linguistic features |  |  |  |  |
| Variable | Description | Type | Unit | Values |
| Posemo | Proportion of words denoting positive emotions in the concatenated transcript | Float | NA | mean 0.05, sd 0.03 |
| Negemo | Proportion of words denoting negative emotions in the concatenated transcript | Float | NA | mean 0.01, sd 0.01 |
| On topic | A score of similarity between the transcripts and the AI description of the image | Float | NA | mean 0.49, sd 0.18 |
| ADJ | Proportion of adjectives in the concatenated transcript | Float | NA | mean 0.04, sd 0.02 |
| PRON | Proportion of pronouns in the concatenated transcript | Float | NA | mean 0.10, sd 0.03 |
| Long words | Proportion of words with more than 7 letters in the concatenated transcript | Float | NA | mean 0.14, sd 0.03 |
| Present verbs | Proportion of verbs in the present tense in the concatenated transcript | Float | NA | mean 0.07, sd 0.04 |
| Past verbs | Proportion of verbs in the past tense in the concatenated transcript | Float | NA | mean 0.03, sd 0.03 |
| Future verbs | Proportion of verbs in the future tense in the concatenated transcript | Float | NA | mean 0.00, sd 0.01 |
| I | Proportion of first-person singular pronouns in the concatenated transcript | Float | NA | mean 0.04, sd 0.03 |
| We | Proportion of first-person plural pronouns in the concatenated transcript | Float | NA | mean 0.00, sd 0.01 |
| You | Proportion of second-person singular pronouns in the concatenated transcript | Float | NA | mean 0.00, sd 0.01 |
| He/she | Proportion of third-person singular pronouns in the concatenated transcript | Float | NA | mean 0.03, sd 0.02 |
| They | Proportion of third-person plural pronouns in the concatenated transcript | Float | NA | mean 0.00, sd 0.01 |
| WC | Number of words in the concatenated transcript | Integer | words | min 72, max 790, median 248 |
| WPS | Median of words per sentence in the concatenated transcript | Float | words/sentence | mean 22.20, sd 15.08 |

**Supplementary Table 2.**
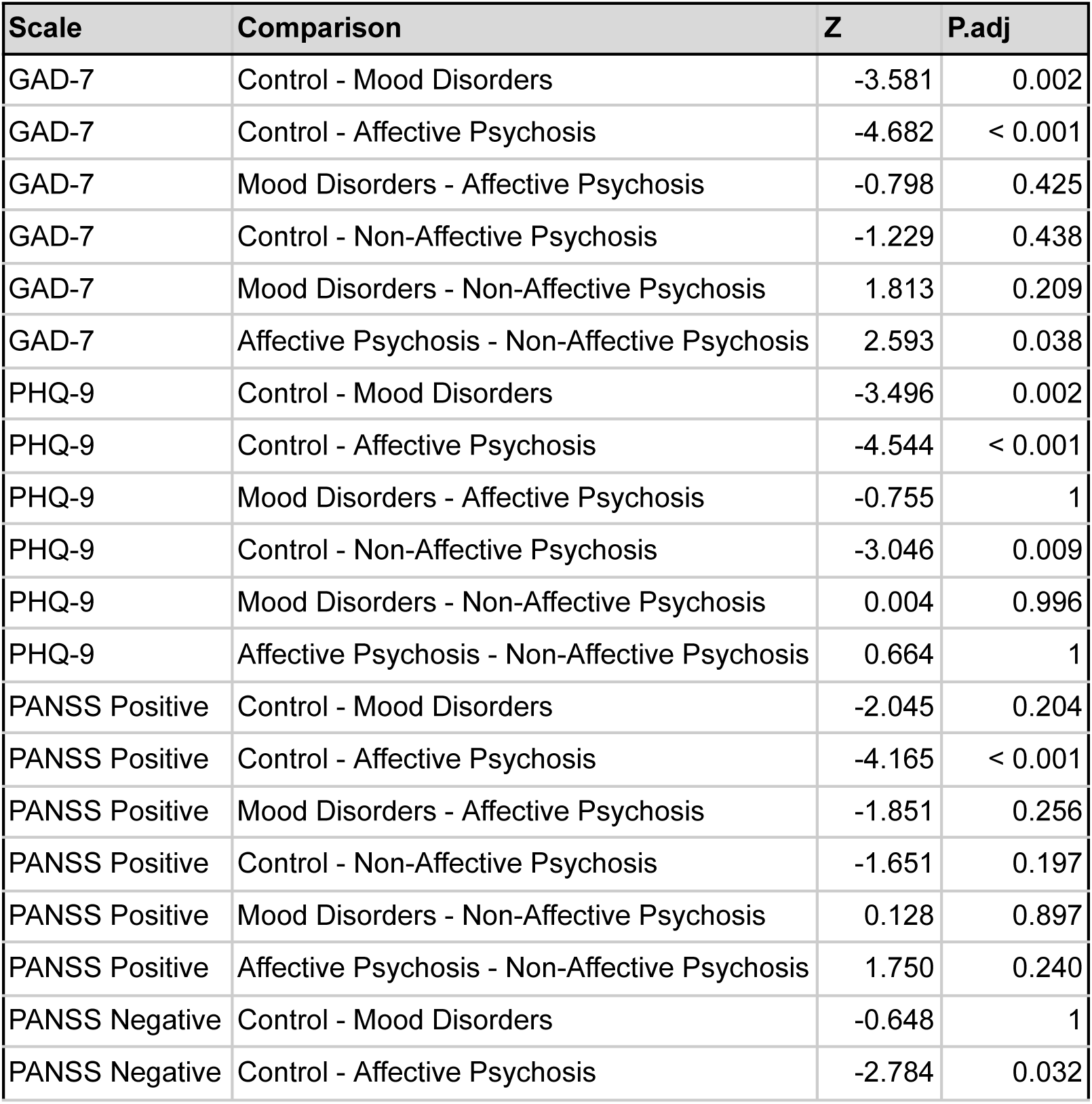
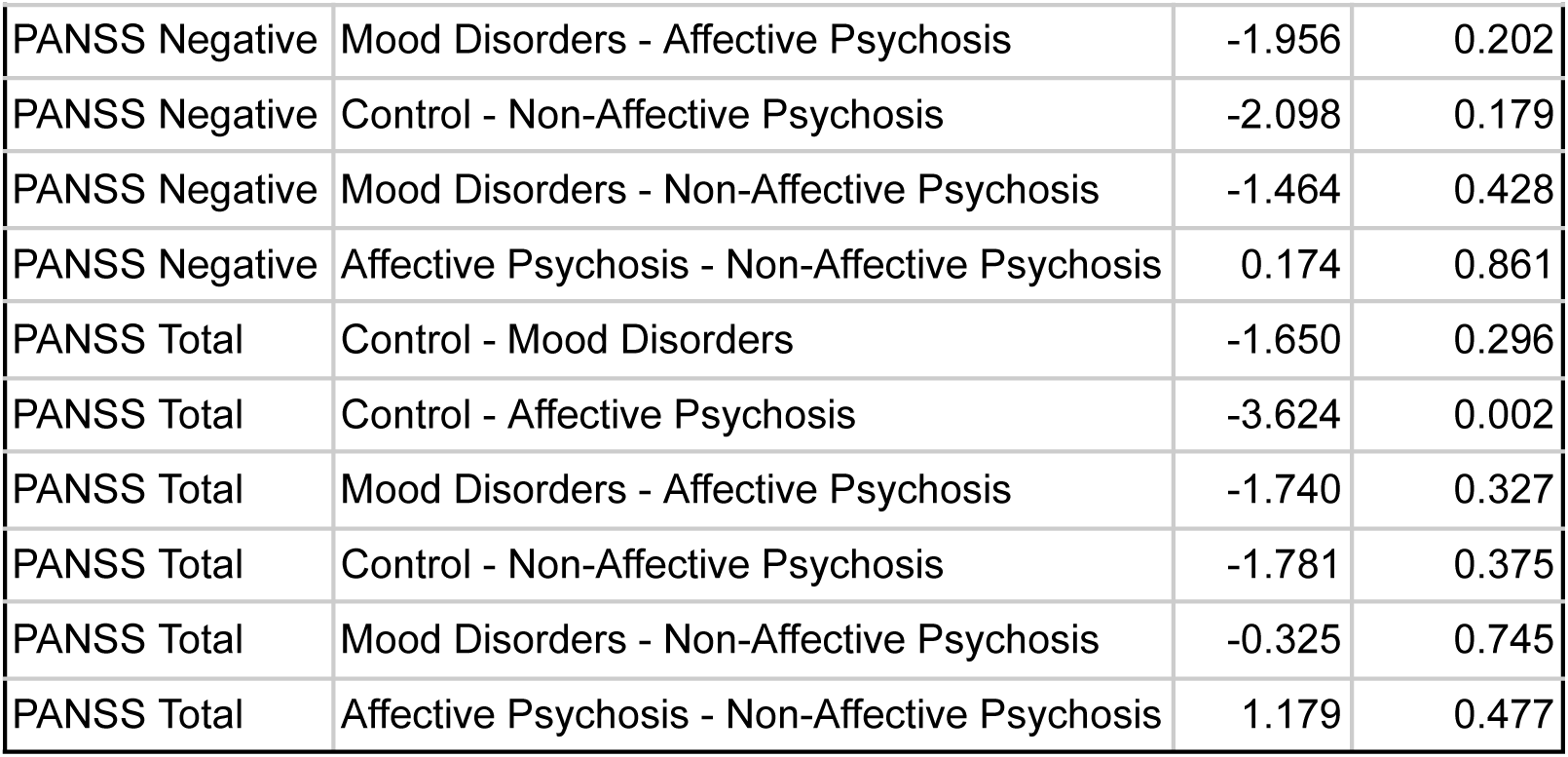
Post hoc pairwise comparisons of model-predicted risk probabilities across clinical diagnostic groups. For each symptom model, posterior mean predicted probabilities of risk were compared across four diagnostic groups: Control (n = 38: 25 healthy controls and 13 university students), Mood Disorders without psychosis (n = 28), Affective Psychosis (n = 36) and Non-Affective Psychosis (n = 18). A Kruskal–Wallis test was followed by Dunn’s pairwise tests with Holm correction for multiple comparisons. Negative Z values mean the first group in each pair had lower predicted probabilities than the second. Models shown are those of Figure 4d: GAD-7 ≥ 10, PHQ-9 ≥ 10, PANSS Positive ≥ 14, PANSS Negative ≥ 14 and PANSS Total ≥ 60. Every model separated the Control and Affective Psychosis groups (adjusted p ≤ 0.032). The GAD-7 and PHQ-9 models also separated Controls from Mood Disorders (adjusted p = 0.002). Only the PHQ-9 model separated Controls from Non-Affective Psychosis (adjusted p = 0.009). P.adj, Holm-adjusted p-value.

